# A diagnostic and economic evaluation of the complex artificial intelligence algorithm aimed to detect 10 pathologies on the chest CT images

**DOI:** 10.1101/2023.04.19.23288584

**Authors:** V. Yu. Chernina, M.G. Belyaev, A.Yu. Silin, I.O. Avetisov, I.A. Pyatnitskiy, E.A. Petrash, M.V. Basova, V.E. Sinitsyn, V.V. Omelyanovsky, V.A. Gombolevskiy

## Abstract

**Background:** Artificial intelligence (AI) technologies can help solve the significant problem of missed findings in radiology studies. An important issue is assessing the economic benefits of implementing AI.

**Aim:** to evaluate the frequency of missed pathologies detection and the economic potential of AI technology for chest CT, validated by expert radiologists, compared with radiologists without access to AI in a private medical center.

**Methods:** An observational, single-center retrospective study was conducted. The study included chest CTs without IV contrast performed from 01.06.2022 to 31.07.2022 in "Yauza Hospital" LLC, Moscow. The CTs were processed using a complex AI algorithm for ten pathologies: pulmonary infiltrates, typical for viral pneumonia (COVID-19 in pandemic conditions); lung nodules; pleural effusion; pulmonary emphysema; thoracic aortic dilatation; pulmonary trunk dilatation; coronary artery calcification; adrenal hyperplasia; osteoporosis (vertebral body height and density changes). Two experts analyzed CTs and compared results with AI. Further routing was determined according to clinical guidelines for all findings initially detected and missed by radiologists. The lost potential revenue (LPR) was calculated for each patient according to the hospital price list.

**Results:** From the final 160 CTs, the AI identified 90 studies (56%) with pathologies, of which 81 studies (51%) were missing at least one pathology in the report. The "second-stage" LPR for all pathologies from 81 patients was RUB 2,847,760 ($37,251 or CNY 256,218). LPR only for those pathologies missed by radiologists but detected by AI was RUB 2,065,360 ($27,017 or CNY 185,824).

**Conclusion:** Using AI for chest CTs as an "assistant" to the radiologist can significantly reduce the number of missed abnormalities. AI usage can bring 3.6 times more benefits compared to the standard model without AI. The use of complex AI for chest CT can be cost-effective.

## Rationale

According to the World Health Organization, most deaths are associated with cardiovascular, oncological, infectious, and lung diseases [1]. Based on large randomized trials of lung cancer screening (LCS), the use of chest low-dose computed tomography (LDCT) in asymptomatic patients at risk resulted in a mortality decrease not only from lung cancer but also from all causes by 6.7% in the National Lung Screening Trial (NLST) and by 39% from year 5 to year 10 of follow-up in the Multicentric Italian Lung Detection (MILD) study, due to the detection of clinically significant incidental findings and the treatment and prevention of related diseases [2,3]. LCS programs have been found to be cost-effective in patient populations at high risk of lung cancer. This effect differs depending on the healthcare systems in different countries [4]. At the same time, it is noted that in these programs, the difference between mortality from lung cancer and total mortality is significant. So, in one of the LCS studies, 77.1% of patients died not from lung cancer but from other causes, such as cardiovascular diseases, other lung diseases, other tumors, infectious diseases, etc. [2]. By focusing on checking for lung cancer, the radiologist may miss pathological findings associated with other conditions. Thus, it was indicated that 58% of clinically significant findings are not reflected in radiologists’ reports in LCS [5].

During the COVID-19 pandemic, LCS programs were suspended as CT scanners were required to perform mass chest CT scans to diagnose this infection. It was noted that in half of the patients who underwent chest CT, incidental findings were detected, and in a quarter, they were clinically significant [6].

The volume of data obtained by CT of the chest allows for diagnosing not only lung pathologies but also other organs and systems diseases [7-9]. Due to a shortage of medical personnel, professional burnout, the effect of a pandemic, and an increase in the doctors’ workload, there is a threat of missing clinically significant findings.

The greatest hopes for solving this problem are placed on artificial intelligence (AI) technologies. An important issue is the assessment of the economic benefits of AI systems. Among AI services for healthcare, the most significant number of products was created for radiology: several times more than in all other medical specialties combined [10]. In the Russian Federation, the largest medical imaging AI assessment project was performed in Moscow. It is the experiment on using innovative technologies in the computer vision field for medical image analysis and further application in the Moscow healthcare system. Within that project, more than 7.5 million radiology studies were processed, including radiographic, mammography, and CT studies [11,12]. Given the preceding, the use of AI algorithms to find a single pathology is of limited value for practical work on fighting critical diseases - the leading causes of death. Given the need for simultaneous detection of several types of pathologies using AI, the first software products that offer a comprehensive chest CT scans analysis have passed all testing stages and are approved for prospective usage in 105 Moscow clinics [13]. One of these products is the complex AI "Multi-IRA" from IRA Labs, which can simultaneously detect ten pathological signs of various diseases on CT [14-17], namely:

1) pulmonary infiltrates, typical for viral pneumonia (COVID-19 in pandemic conditions) (U07 according to the international classification of diseases of the 10th revision (ICD-10)) with an assessment of the lung damage percentage;

2) lung nodules with an assessment of size, volume, and density to detect malignant neoplasms in the lungs (C34 according to ICD-10);

3) pleural effusion (J94 according to ICD-10);

4) pulmonary emphysema as a manifestation of COPD (J44 according to ICD-10);

5) thoracic aorta diameter measurement to detect its dilatation and aneurysms (I70 and I71 according to ICD-10);

6) pulmonary trunk diameter measurement to detect the causes of possible pulmonary hypertension (I27 according to ICD-10);

7) analysis of coronary artery calcification severity according to the Agatston index to assess coronary atherosclerosis and the risk of coronary heart disease (I20-I25 according to ICD-10);

8) assessment of adrenal glands thickness to look for masses and hyperplasia (C74 according to ICD-10);

9) vertebral body heights for diagnosing compression fractures (M80-M85 according to ICD-10);

10) vertebral bodies density analysis to detect osteoporosis (M80-M85 according to ICD-10).

## Purpose of the study

To evaluate the frequency of detection of significant pathological findings and the economic potential of using complex AI technologies in the chest CT scans analysis with radiologists’ verification, compared with radiologists without access to AI in a private medical center.

## Methods

### Study design

An observational single-center retrospective study was carried out. Informed consent from patients was not required. The paper was prepared following the CHEERS 2022 checklist, designed for the economic evaluation of medical research [18,19]. An economic analysis plan was developed to assess the lost potential revenue (LPR) that should be provided to patients according to clinical guidelines and best practices of evidence-based medicine based on pathological findings. The use of complex AI can bring LPR beyond what radiologists get through further diagnostic actions to clarify the nature and severity of CT findings.

The scheme of the study is shown in Figure 1.

**Figure 1.**
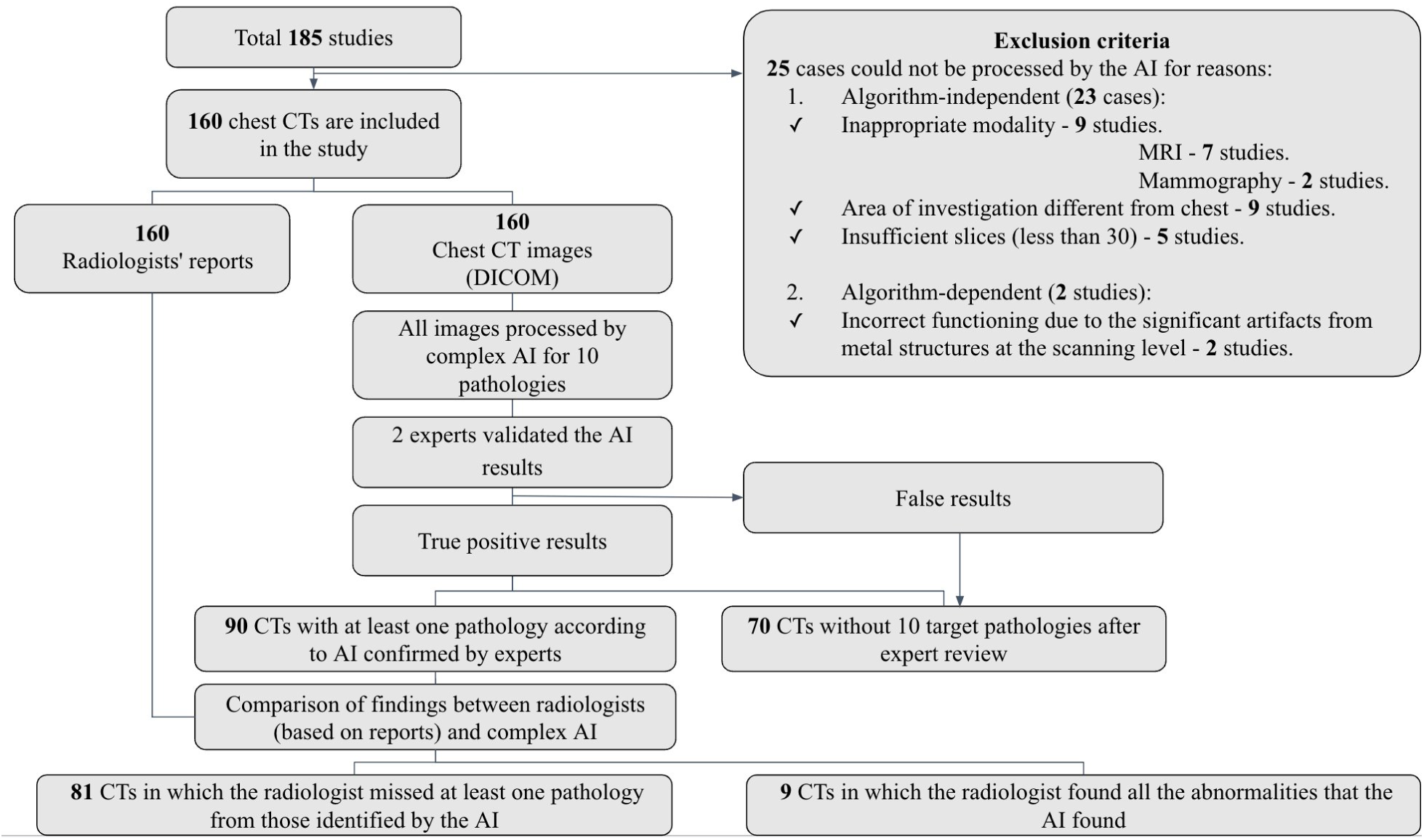
Study design.

### Compliance Criteria

Inclusion and exclusion criteria were used to form the study group.

#### Inclusion Criteria

- Chest CT scans of men and women who applied for medical services in the private Moscow clinic;
- Chest CT scans were performed and interpreted by radiologists in the period from 06/01/2022 to 07/31/2022;
- Non-contrast Chest CT scans;
- Age over 18;
- Availability of chest CT images in DICOM format and radiology reports;
- The patient’s first visit to a healthcare facility.

#### Exclusion Criteria

- Age over 85;
- The previous chest CT scan was performed within one year;
- The AI could not process the study due to reasons beyond the algorithm’s control, such as inappropriate modality, study area other than the chest, and an insufficient number of slices (less than 30).
- AI could not process the study for reasons that depend on the algorithm’s peculiarities: for example, incorrect work due to the presence of pronounced artifacts from metal structures at the scanning level.

### Terms and Conditions

CT studies were performed in a multidisciplinary private clinic, "Yauza Hospital" LLC, which provides primary health care and specialized medical care to the adult population in Moscow.

### Study duration

The study was conducted on CT data performed in the period from 06/01/2022 to 07/31/2022. A retrospective analysis using AI and verification of the results by experts were carried out from 10/01/2022 to 11/30/2022.

## Acquisition technique and Image analysis

Non-contrast chest CT scans were performed on a Philips Ingenuity 128-Slice CT scanner. The chest scanning protocol followed standard equipment manufacturer recommendations and national guidelines. CT results were provided to physicians and AI in 2 series: performed during reconstruction with a soft tissue kernel (60 HU - window center, 360 HU - window width) and a lung kernel (-500 HU - window center, 1500 HU - window width). The slice thickness was 1.0 mm. Iterative model reconstruction (IMR) algorithms were used to improve image quality (noise reduction) and reduce the radiation exposure dose to the patient.

All CT scans were processed using the IRA Labs complex AI program "Multi-IRA", integrated into the clinic’s electronic archive, Picture Archiving and Communication System Radiology information systems (PACS-RIS). The AI algorithms used in this study were previously tested on specially prepared calibration datasets as part of the Moscow Experiment on the Application of AI products (ClinicalTrials.gov Identifier: NCT04489992)[20-23].

The criterion for AI algorithm usage possibility was the accuracy not lower than the area under the ROC curve (ROC AUC) of 0.81 for each pathology, according to the guidelines for clinical trials of software based on intelligent technologies [24]. The values of diagnostic accuracy metrics for AI algorithms, obtained on developer-independent non-public datasets within the framework of the Moscow Experiment, are presented in Table 1 [11-13].

**Table 1.** Values of diagnostic accuracy metrics for chest CT complex AI derived from datasets in the Moscow Experiment.

| No | Name of the AI algorithm<br>"Multi-IRA" for selected<br>pathologies | ROC<br>AUC | Sensitivity | Specificity | Accuracy |
| --- | --- | --- | --- | --- | --- |
| 1 | COVID-IRA<br>(pulmonary infiltrates<br>detection) | 0,98 | 0,95 | 0,94 | 0,94 |
| 2 | LungNodule -IRA<br>(lung nodules detection) | 0,932 | 0,86 | 0,9 | 0,88 |
| 3 | PleuralEffusion-IRA<br>(pleural effusion detection ) | 0,999 | 0,98 | 1 | 0,99 |
| 4 | Aorta-IRA (chest)<br>(thoracic aortic dilatation<br>detection) | 0,997 | 0,96 | 1 | 0,98 |
| 5 | Aorta-IRA (chest +<br>abdomen)<br>(thoracic and abdominal<br>aortic dilatation detection) | 1 | 0,98 | 1 | 0,99 |
| 6 | PulmTrunk-IRA<br>(pulmonary trunk dilatation<br>detection) | 1 | 1 | 0,98 | 0,99 |
| 7 | Agatston-IRA<br>(coronary artery<br>calcification detection) | 0,986 | 0,96 | 0,96 | 0,96 |
| 8 | Genant-IRA<br>(osteoporosis detection) | 0,995 | 1 | 0,98 | 0,99 |
| 9 | Emphysema-IRA<br>(emphysema detection) | 0,989 | 0,94 | 0,98 | 0,96 |
| 10 | Adrenal-IRA<br>(adrenal masses and<br>hyperplasia detection) | 0,96 | 1 | 0,96 | 0,98 |

## The primary outcome of the study

For all the findings identified or missed by the radiologists, “second stages” were determined, i.e., further patient routing following current clinical guidelines for each disease. They were: consultations with specialized specialists and various clinical, instrumental, and laboratory additional examinations. This study did not evaluate the cost of treatment.

Then, for each patient, the LPR was calculated according to the price list of the clinic, which was determined based on the necessary but not delivered medical services following the clinical recommendations for missed pathologies. Also, LPR was calculated only for significant missed pathologies. A detailed classification of significant and non-significant pathologies is presented in Table 2. The absence of information about a pathological finding in the radiology reports in the electronic medical information system was considered missed diagnostic findings when detected by an AI algorithm and verified by an expert radiologist. Ground truth was defined as findings verified by two expert radiologists.

**Table 2.**
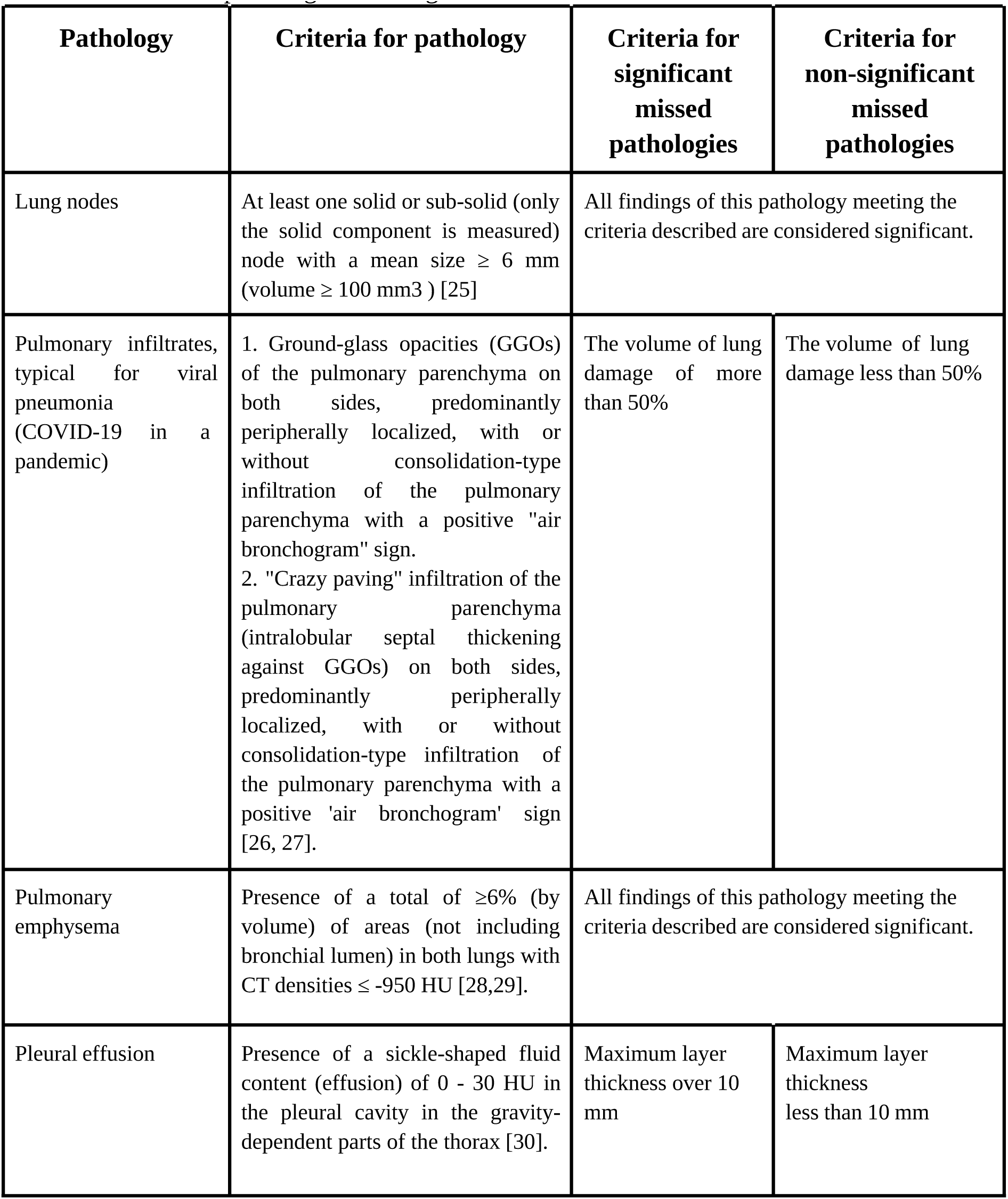
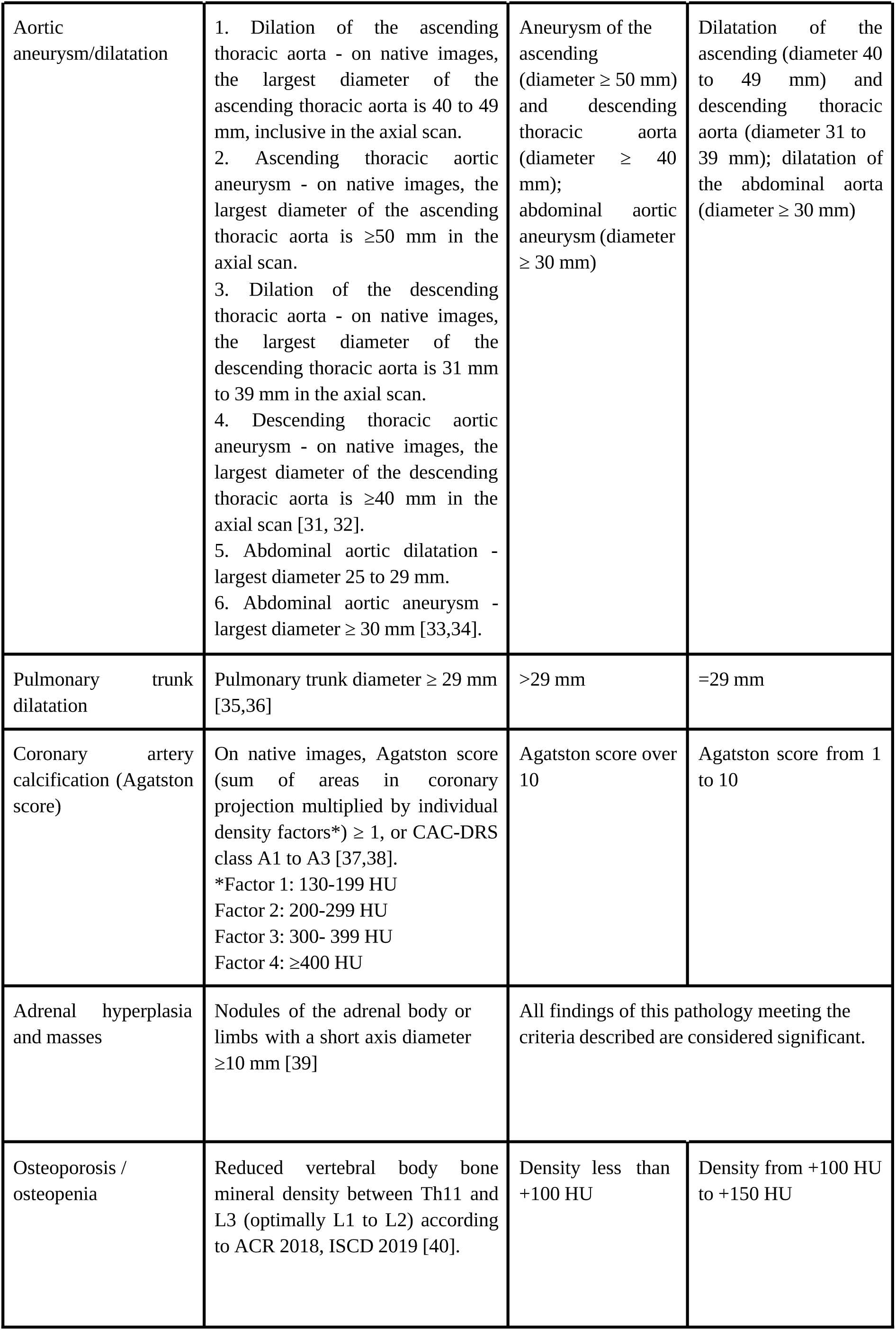
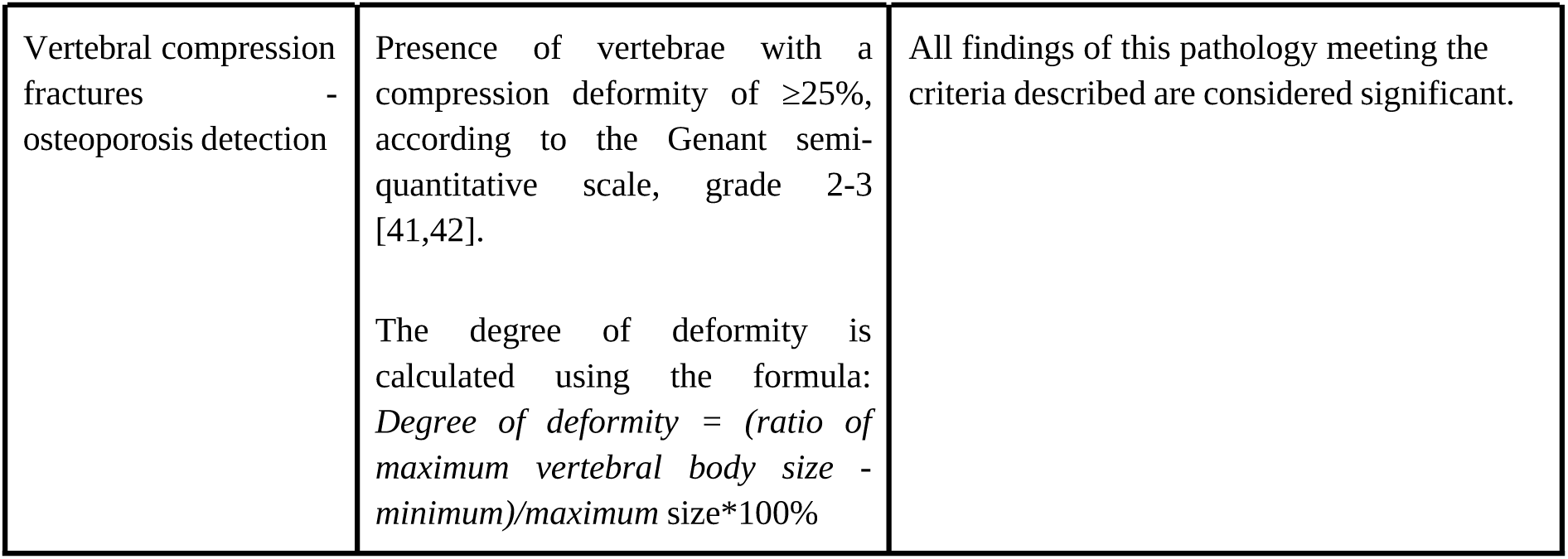
Criteria for pathological findings.

| **Pathology** | **Criteria for pathology** | **Criteria for significant missed pathologies** | **Criteria for non-significant missed pathologies** |
| --- | --- | --- | --- |
| Lung nodes | At least one solid or sub-solid (only the solid component is measured) node with a mean size $\geq 6$ mm (volume $\geq 100$ mm<sup>3</sup>) [25] | All findings of this pathology meeting the criteria described are considered significant. | |
| Pulmonary infiltrates, typical for viral pneumonia (COVID-19 in a pandemic) | 1. Ground-glass opacities (GGOs) of the pulmonary parenchyma on both sides, predominantly peripherally localized, with or without consolidation-type infiltration of the pulmonary parenchyma with a positive "air bronchogram" sign.<br>2. "Crazy paving" infiltration of the pulmonary parenchyma (intralobular septal thickening against GGOs) on both sides, predominantly peripherally localized, with or without consolidation-type infiltration of the pulmonary parenchyma with a positive 'air bronchogram' sign [26, 27]. | The volume of lung damage of more than 50% | The volume of lung damage less than 50% |
| Pulmonary emphysema | Presence of a total of $\geq 6\%$ (by volume) of areas (not including bronchial lumen) in both lungs with CT densities $\leq -950$ HU [28,29]. | All findings of this pathology meeting the criteria described are considered significant. | |
| Pleural effusion | Presence of a sickle-shaped fluid content (effusion) of 0 - 30 HU in the pleural cavity in the gravity-dependent parts of the thorax [30]. | Maximum layer thickness over 10 mm | Maximum layer thickness less than 10 mm |
| Aortic aneurysm/dilatation | 1. Dilation of the ascending thoracic aorta - on native images, the largest diameter of the ascending thoracic aorta is 40 to 49 mm, inclusive in the axial scan.<br>2. Ascending thoracic aortic aneurysm - on native images, the largest diameter of the ascending thoracic aorta is $\geq 50$ mm in the axial scan.<br>3. Dilation of the descending thoracic aorta - on native images, the largest diameter of the descending thoracic aorta is 31 mm to 39 mm in the axial scan.<br>4. Descending thoracic aortic aneurysm - on native images, the largest diameter of the descending thoracic aorta is $\geq 40$ mm in the axial scan [31, 32].<br>5. Abdominal aortic dilatation - largest diameter 25 to 29 mm.<br>6. Abdominal aortic aneurysm - largest diameter $\geq 30$ mm [33,34]. | Aneurysm of the ascending (diameter $\geq 50$ mm) and descending thoracic aorta (diameter $\geq 40$ mm); abdominal aortic aneurysm (diameter $\geq 30$ mm) | Dilatation of the ascending (diameter 40 to 49 mm) and descending thoracic aorta (diameter 31 to 39 mm); dilatation of the abdominal aorta (diameter $\geq 30$ mm) |
| Pulmonary trunk dilatation | Pulmonary trunk diameter $\geq 29$ mm [35,36] | $>29$ mm | $=29$ mm |
| Coronary artery calcification (Agatston score) | On native images, Agatston score (sum of areas in coronary projection multiplied by individual density factors*) $\geq 1$, or CAC-DRS class A1 to A3 [37,38].<br>*Factor 1: 130-199 HU<br>Factor 2: 200-299 HU<br>Factor 3: 300- 399 HU<br>Factor 4: $\geq 400$ HU | Agatston score over 10 | Agatston score from 1 to 10 |
| Adrenal hyperplasia and masses | Nodules of the adrenal body or limbs with a short axis diameter $\geq 10$ mm [39] | All findings of this pathology meeting the criteria described are considered significant. | |
| Osteoporosis / osteopenia | Reduced vertebral body bone mineral density between Th11 and L3 (optimally L1 to L2) according to ACR 2018, ISCD 2019 [40]. | Density less than +100 HU | Density from +100 HU to +150 HU |

|  |  |  |
| --- | --- | --- |
| Vertebral compression fractures - osteoporosis detection | Presence of vertebrae with a compression deformity of $\geq 25\%$, according to the Genant semi-quantitative scale, grade 2-3 [41,42].<br>The degree of deformity is calculated using the formula:<br>*Degree of deformity = (ratio of maximum vertebral body size - minimum)/maximum size\*100%* | All findings of this pathology meeting the criteria described are considered significant. |

## Additional study outcomes

Additionally, the number and percentages of reports with significant and insignificant errors were calculated for each radiologist.

## Subgroup Analysis

### Outcome Registration Methods

Two experts (radiologists with 10 and 13 years of experience, not employees of the clinic participating in this study) reviewed the CT scan along with AI algorithm results to eliminate false positive results. With opposing opinions, a single decision was made after a collegiate discussion. As a result of this analysis, the true positive AI results, confirmed by two experts, were selected. Then, pathological findings detected by AI were compared with initial radiology protocols, and cases of missing pathologies were identified. All the missed pathologies were divided into significant and insignificant. The criteria for the pathologies’ significance were determined following the requirements by the scientific and problem commission of the Scientific and Practical Clinical Center for Diagnostics and Telemedicine Technologies within the Moscow Experiment (protocols dated 10.12.2021 No. 9/2021, dated 28.02.2022 No. 1/ 2022, dated 06.12.2022 No. 7/2022, dated 13.01.2023 No. 1/2023). These requirements are based on clinical guidelines and evidence-based best practices. This study did not evaluate the presence/absence of epicardial fat in the radiology protocols since this clinic’s radiologists did not have tools to measure the volume of adipose tissue. The criteria for pathologies and the distribution by the significance of missed findings are presented in Table 2.

## Ethical expertise

A notification was sent to the Independent Ethical Committee of the Moscow Regional Branch of the Russian Society of Radiologists about a retrospective study (protocol dated 03/01/2023).

## Statistical analysis

Methods of statistical data analysis:

Descriptive statistics methods were used to present the results, indicating the absolute number (n) and percentage (%) of observations in each category. A comparison of the detection rates of pathologies by different methods was carried out using the Z-test for proportions. The p-values obtained for each of the nine pathologies were corrected for multiple testing (within the framework of the general hypothesis of the absence of a statistically significant difference between the diagnostic results) by the Bonferroni correction. Analysis of financial performance was performed using a paired t-test. The level of statistical significance for p-values was considered to be 0.05. Statistical analysis was carried out using the R v program. 4.1.3.

## Results

Objects (participants) of the study

A total of 185 native chest CT scans were selected (male/female ratio 47/53%, age from 19 to 83 years, mean age 49.5 years) that met the specified criteria. Of these, 25 studies could not be processed by AI for the following reasons:

1. AI-independent AI (23 studies):
  ✓ Inappropriate modality - 9 studies.
    - 7 examinations.
    - 2 examinations.
  ✓ Non-chest CT - 9 studies.
  ✓ The insufficient number of slices (less than 30), incl. localizers - 5 studies.
2. AI-dependent (2 studies):
  ✓ Incorrect work due to pronounced artifacts from metal structures at the scanning level - 2 studies.

The final group for analysis consisted of 160 chest CT studies with radiology reports. Additional information on the presence of oncological, cardiovascular, and other chronic diseases in patients was not collected since patients applied to this clinic only under compulsory medical insurance and voluntary medical insurance policies or on a paid basis to specific specialists. The health data in this clinic’s medical information system was most likely incomplete.

## Primary results of the study

Automatic anonymization and transfer of CT studies from the clinic to the complex AI developer were set up. Then the AI results were returned to the clinic and expert radiologists for validation. The experts provided a list of all discrepancies between the verified AI results and the initial radiology reports for quality control (Figure 2). There were no appeals from the clinic. The largest number of clinically significant missed findings was found in cases of osteoporosis and adrenal masses (14 each). The biggest number of insignificant pathologies was found for aortic dilatation (36 cases) and osteopenia (40 cases). Detailed results by the number of findings are shown in Figure 3.

**Figure 2.**
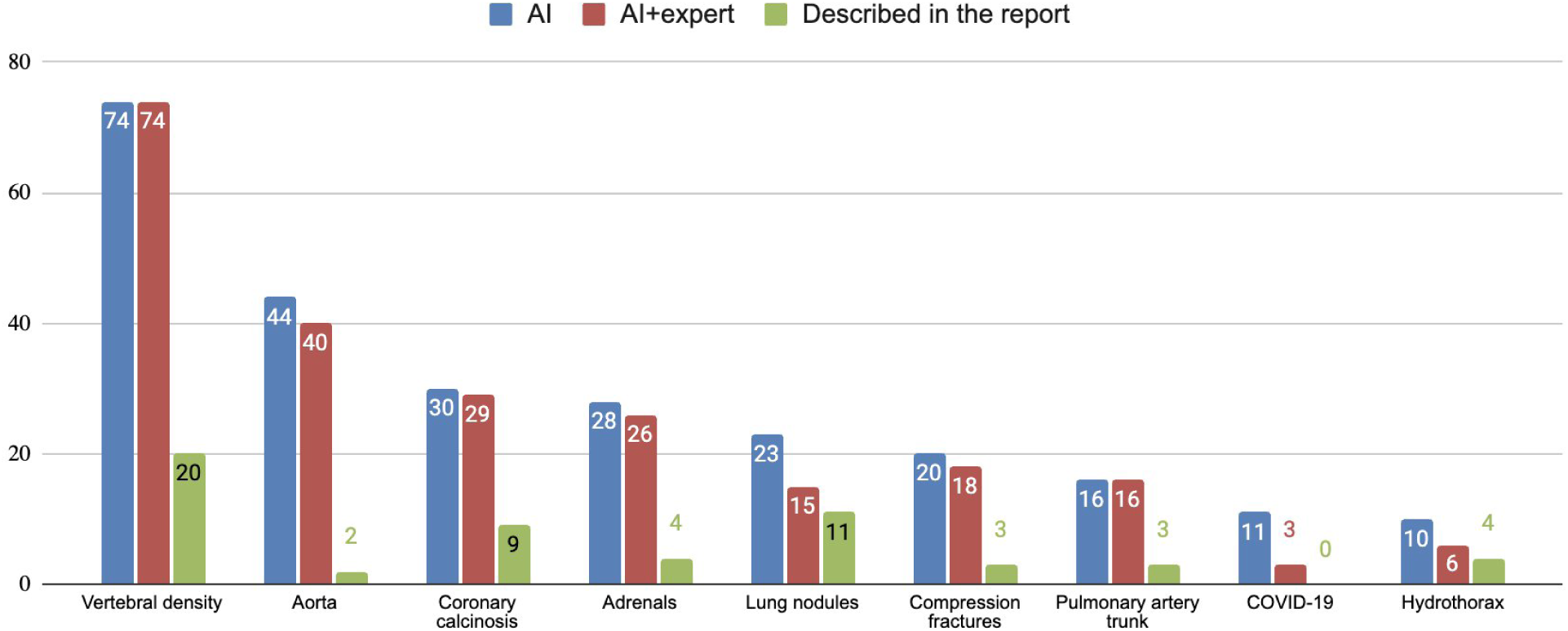
Results by the number of findings detected with and without AI.

**Figure 3.**
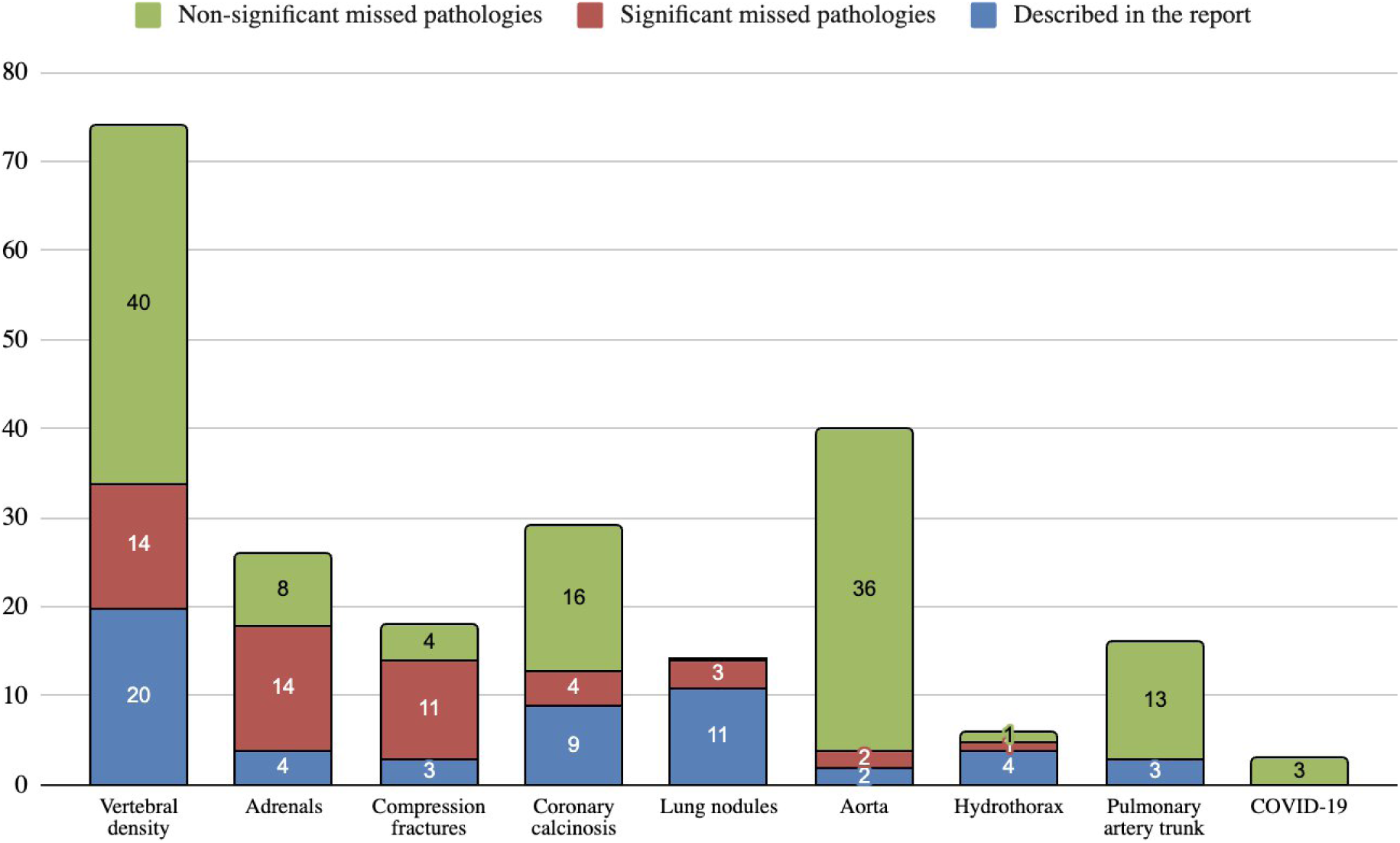
Results by the number of findings (ranked by the number of significant missed findings).

With the help of AI, 90 (56%) studies with pathologies were identified, of which at least one pathology was missed in 81 (51%) radiology reports. In 70 studies, the AI algorithm did not reveal any pathology. It should be noted that there may have been other pathological findings in the presented studies that were not included in this study’s complex AI program analysis. A summary of the analysis is shown in Table 3.

**Table 3:** Summary of the number of patients with detected and missed pathologies.

|  | Quantity | % of all cases | Estimated % of abnormalities |
| --- | --- | --- | --- |
| Total number of patients | 160 | 100% | - |
| No pathological findings | 70 | 44% | - |
| With pathological findings | 90 | 56% | 100% |
| Radiologist has found at least one pathology among those detected by the AI | 35 | 22% | 39% |
| Radiologist has missed at least one pathology among those detected by the AI | 81 | 51% | 90% |
| CT studies where only the AI algorithm found pathological changes | 55 | 34% | 61% |

In one CT study, there could be several pathologies, some of which were described by a radiologist in the report, and the other part was detected only by AI (potential benefit of AI).

A detailed presentation of the analysis by LPR due to missed pathologies for each of the 90 patients is shown in Figure 4.

**Figure 4.**
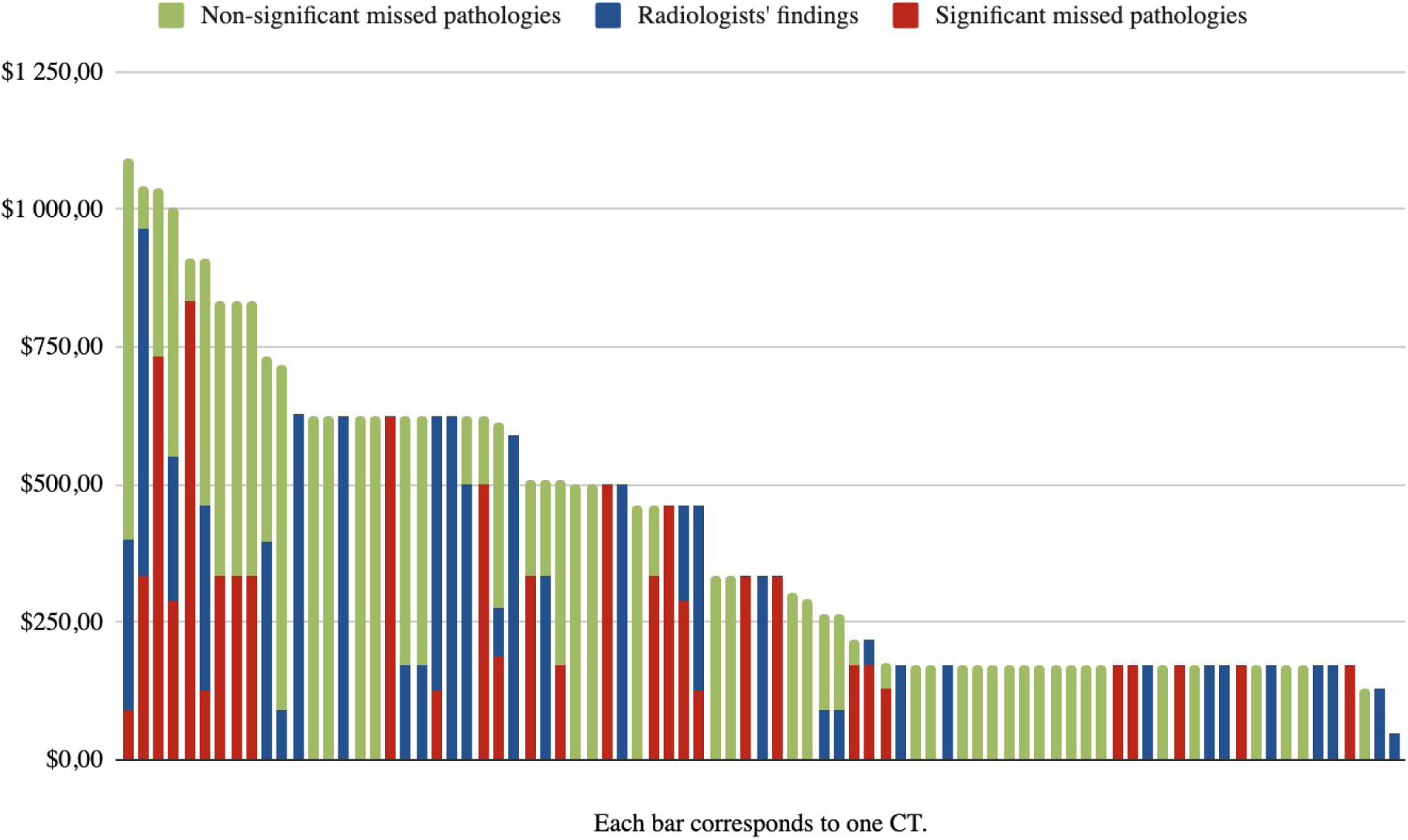
Lost potential revenue (LPR) analysis due to missed pathologies on CT scans.

Algorithmic analysis approaches with expert verification against the clinic’s radiologist without AI were statistically significant for the following types of pathological findings: aortic aneurysms/dilations, pulmonary trunk dilation, assessment of the coronary artery calcification score, vertebral compression fractures, decreased density of the vertebral bodies and thickening of the adrenal glands (Table 4).

**Table 4.** Comparison of the two diagnostic approaches pathologies detection.

| Parameter | Number of cases detected | Proportion [95% CI] | 95% CI for the difference in fractions | p-value Z-test H0: fractions are equal (Bonferroni correction) |
| --- | --- | --- | --- | --- |
| **COVID-19 signs** |  |  |  |  |
| AI Algorithm + Expert | 3 | 0.016<br>[0.003; 0.047] | [-0,008; 0,04] | 0,246 (1) |
| Radiology report | 0 | 0 [0; 0,02] |  |  |
| **Lung nodes** |  |  |  |  |
| AI Algorithm + Expert | 15 | 0.082<br>[0.047; 0.132] | [-0,036 0,08] | 0,542 (1) |
| Radiology report | 11 | 0,06 [0,031; 0,106] |  |  |
| **Pleural effusion** |  |  |  |  |
| AI Algorithm + Expert | 6 | 0,033 [0,012; 0,07] | [-0,028; 0,05] | 0,749 (1) |
| Radiology report | 4 | 0.022<br>[0.006; 0.055] |  |  |
| **Aorta dilatation** |  |  |  |  |
| AI Algorithm + Expert | 40 | 0,22 [0,162; 0,287] | [0,141; 0,276] | < 0,001 (< 0,001) |
| Radiology report | 2 | 0.011<br>[0.001; 0.039] |  |  |
| **Pulmonary trunk dilatation** |  |  |  |  |
| AI Algorithm + Expert | 16 | 0.088<br>[0.051; 0.139] | [0,021; 0,122] | 0,005 (0,042) |
| Radiology report | 3 | 0.016<br>[0.003; 0.047] |  |  |
| **Coronary artery calcification** |  |  |  |  |
| AI Algorithm + Expert | 29 | 0.159<br>[0.109; 0.221] | [0,043; 0,177] | 0,001 (0,01) |
| Radiology report | 9 | 0.049<br>[0.023; 0.092] |  |  |
| **Compression vertebrae fractures** |  |  |  |  |
| AI Algorithm + Expert | 18 | 0,1 [0,06; 0,152] | [0,03; 0,135] | 0,002 (0,015) |
| Radiology report | 3 | 0.016<br>[0.003; 0.047] |  |  |
| **Evaluation of vertebral body mineral density** |  |  |  |  |
| AI Algorithm + Expert | 74 | 0.407<br>[0.335; 0.482] | [0,207; 0,387] | < 0,001 (< 0,001) |
| Radiology report | 20 | 0,11 [0,068; 0,165] |  |  |
| **Adrenal thickening** |  |  |  |  |
| AI Algorithm + Expert | 26 | 0.143<br>[0.095; 0.202] | [0,06; 0,181] | < 0,001 (0,001) |
| Radiology report | 4 | 0.022<br>[0.006; 0.055] |  |  |

Estimated "Stage 2" LPR for all missed pathologies in 81 patients totaled RUB 2,847,760 ($37,251 or CNY 256,218), or RUB 17,799 per patient ($233 or CNY 1,601). The "second stage" LPR only for those pathologies that were missed in the radiology reports but were identified by AI (and confirmed by experts) totaled RUB 2,065,360 ($27,017 or CNY 185,824), or RUB 12,909 per patient ($169 or CNY 1,161). The results of the LPR calculation for all findings are presented in Table 5.

**Table 5.** Analysis of estimated LPR from all missed CT findings.

|  | Calculation for all patients<br>(USD/Yuan) |  |  |  | Calculation per patient<br>(USD/Yuan) |  |  |  |
| --- | --- | --- | --- | --- | --- | --- | --- | --- |
|  | Total<br>cost<br>(all) | Consult.<br>(all) cost | Pre-exami<br>nations<br>(all) cost | Follow<br>- u p<br>(all)<br>cost | Total<br>cost<br>(all) | Counseling<br>(all) cost | Pre-examina<br>tions (all)<br>cost | Follow<br>- u p<br>(all)<br>cost |
| Cost of<br>'second<br>stage'<br>diagnosis<br>for all<br>pathologie<br>s | 37<br>250,99<br>/<br>256<br>217,95 | 6 060,34/<br>41 683,91 | 26 812,51/<br>184 420,49 | 4<br>378,15/<br>30<br>113,54 | 232,83/<br>1<br>601,41 | 37,88/<br>260,56 | 167,58/<br>1 152,63 | 27,37/<br>188,22 |
| The cost of<br>a 'second<br>stage' for<br>only those<br>pathologie<br>s missed<br>by the<br>doctor and<br>found by<br>the AI<br>algorithm | 27<br>016,57<br>/<br>185<br>824,05 | 4 274,81/<br>29 402,77 | 19 875,76/<br>136 708,47 | 2 866/<br>19<br>712,81 | 168,86/<br>1<br>161,45 | 26,72/<br>183,81 | 124,23/<br>854,46 | 17,91/<br>123,17 |
| The cost of<br>a "second<br>stage" for<br>pathologie<br>s found by<br>radiologist | 10<br>234,42<br>/<br>70<br>393,9 | 1 785,53/<br>12 281,14 | 6 936,75/<br>47 712,02 | 1<br>512,14/<br>10<br>400,73 | 63,97/<br>439,96 | 11,16/<br>76,75 | 43,35/<br>298,17 | 9,46/<br>65,05 |

According to the results, the total LPR only from significant missed pathologies was RUB 770,855 ($10,083 or CNY 69,355), or RUB 4,818 per patient ($63 or CNY 433). The analysis results are presented in Table 6.

**Table 6.**
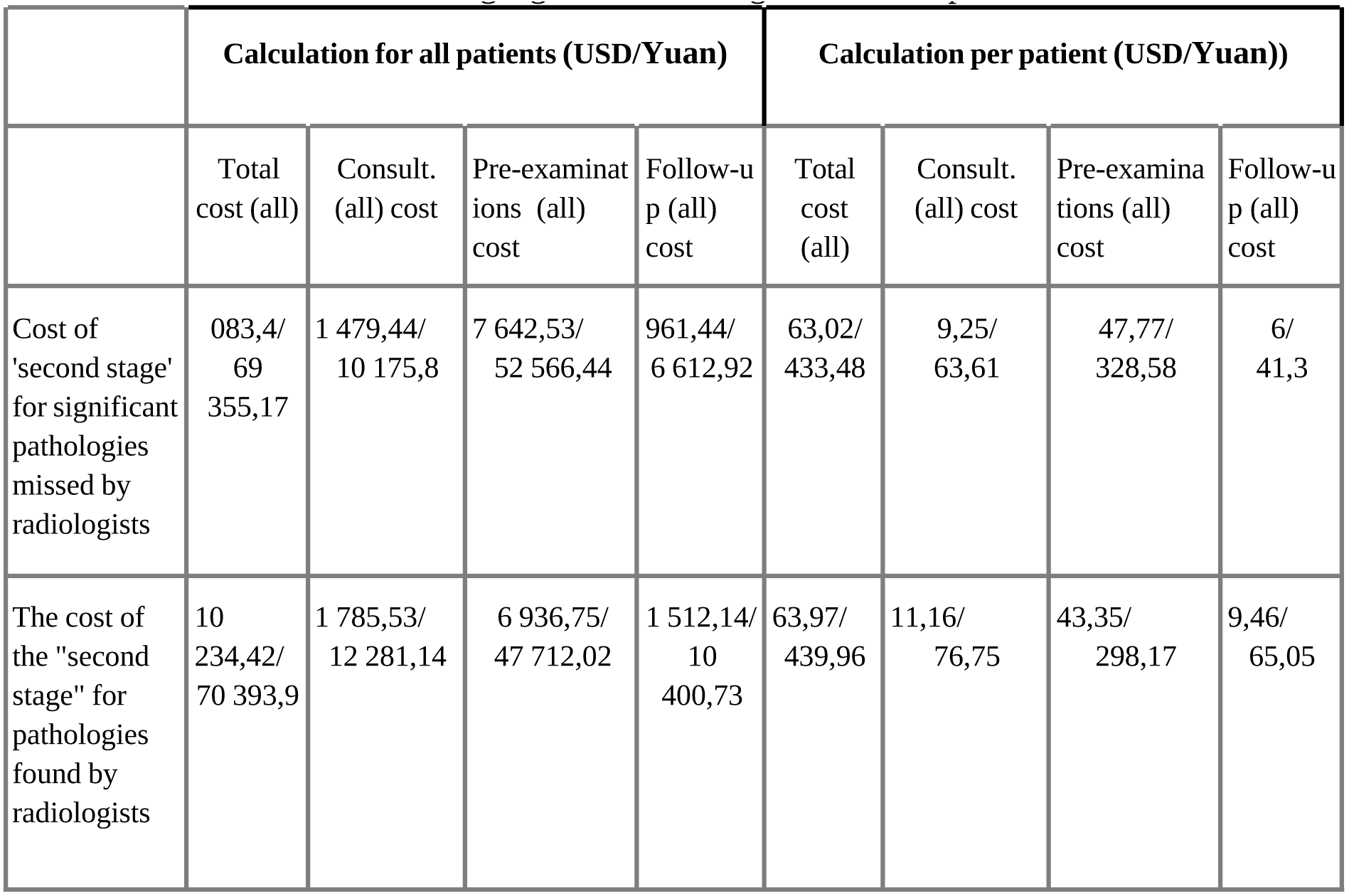
LPR results from missing significant findings. *LPR - lost potential revenue*.

|  | Calculation for all patients (USD/Yuan) |  |  |  | Calculation per patient (USD/Yuan) |  |  |  |
| --- | --- | --- | --- | --- | --- | --- | --- | --- |
|  | Total cost (all) | Consult. (all) cost | Pre-examinations (all) cost | Follow-up (all) cost | Total cost (all) | Consult. (all) cost | Pre-examinations (all) cost | Follow-up (all) cost |
| Cost of 'second stage' for significant pathologies missed by radiologists | 083,4/69<br>355,17 | 1 479,44/<br>10 175,8 | 7 642,53/<br>52 566,44 | 961,44/<br>6 612,92 | 63,02/<br>433,48 | 9,25/<br>63,61 | 47,77/<br>328,58 | 6/<br>41,3 |
| The cost of the "second stage" for pathologies found by radiologists | 10<br>234,42/<br>70 393,9 | 1 785,53/<br>12 281,14 | 6 936,75/<br>47 712,02 | 1 512,14/<br>10<br>400,73 | 63,97/<br>439,96 | 11,16/<br>76,75 | 43,35/<br>298,17 | 9,46/<br>65,05 |

Costs were compared using a paired t-test, calculating the mean difference per patient and constructing a 95% confidence interval. Thus, 160*12908.5 [160* 9833.5; 160*15983.5] gives the total LPR for the analysis population with its own confidence interval (CI). The results are presented in Table 7.

**Table 7.**
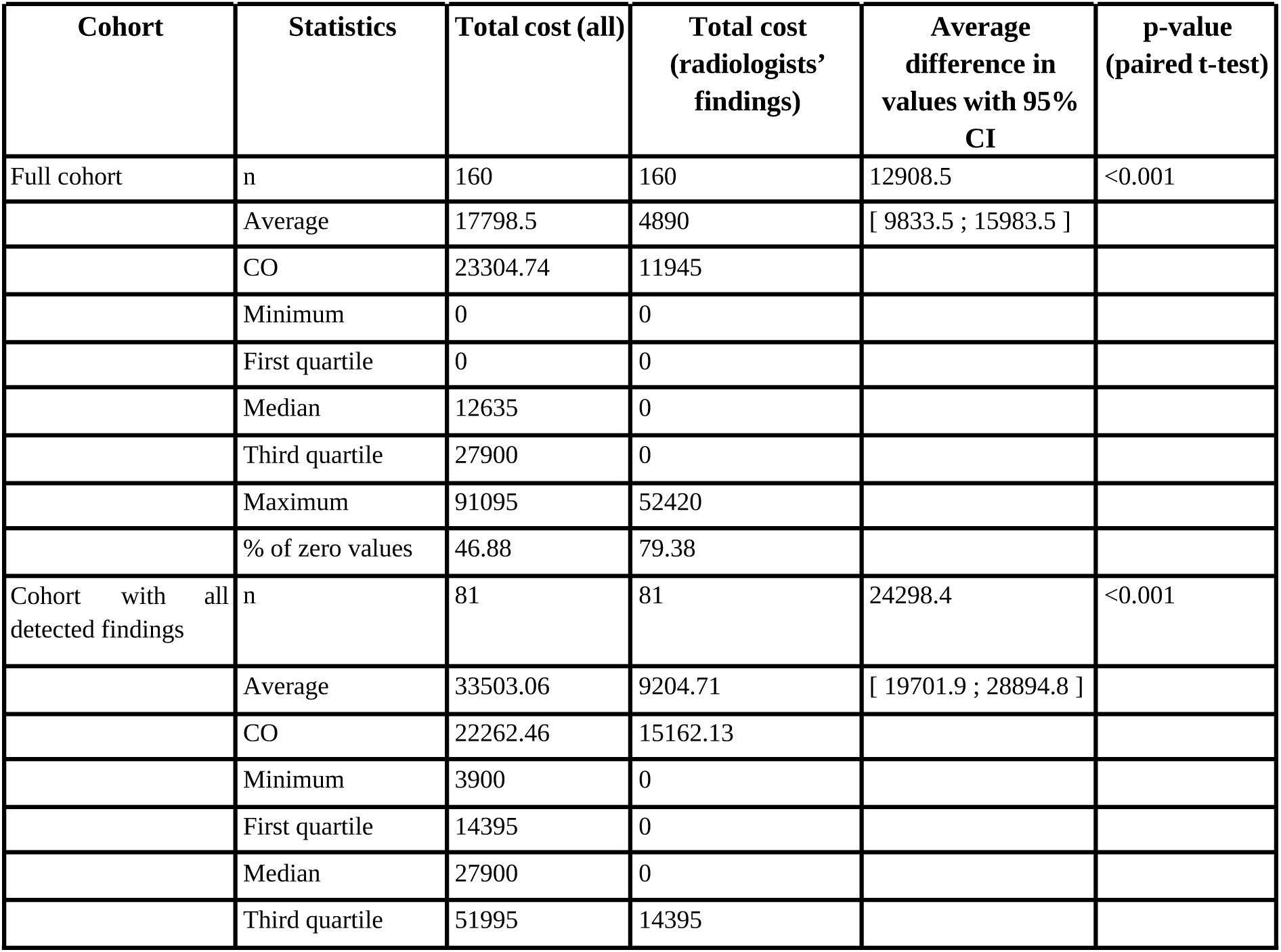
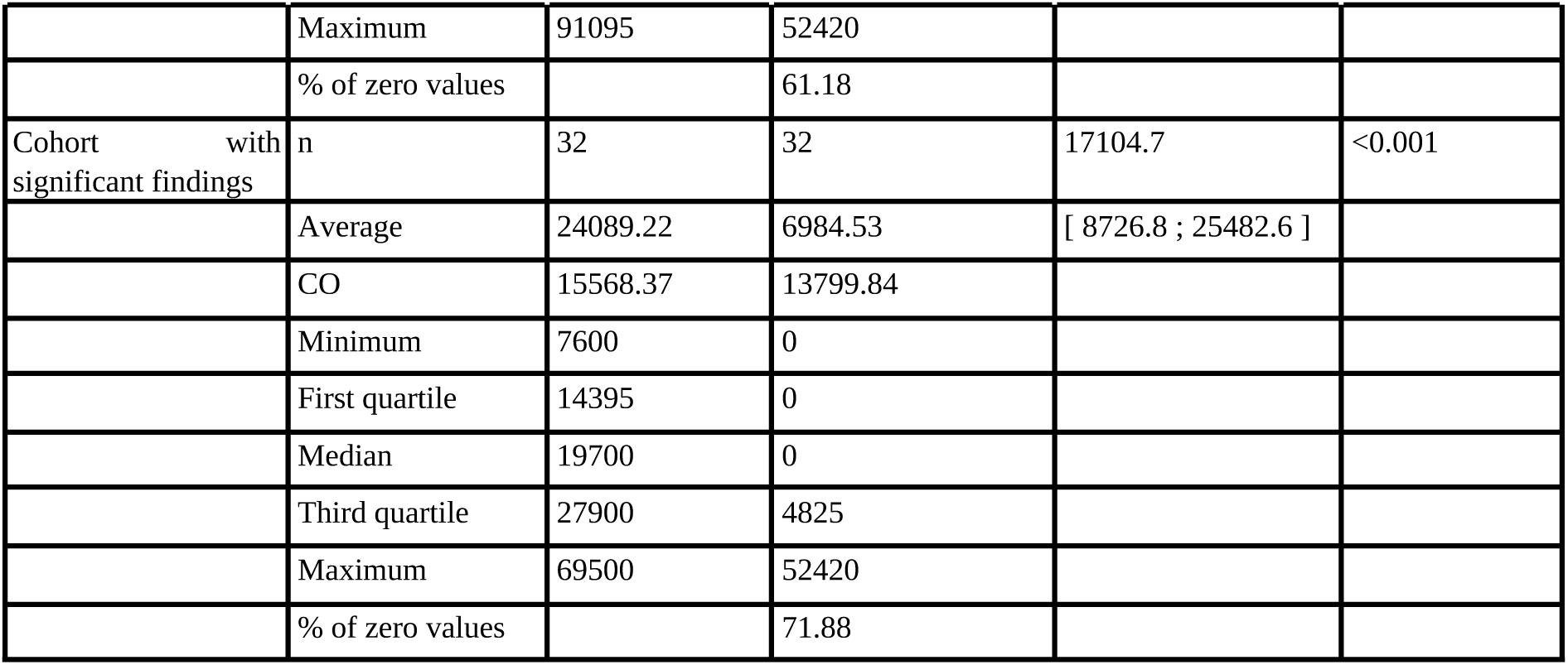
Cost-effectiveness.

| Cohort | Statistics | Total cost (all) | Total cost (radiologists' findings) | Average difference in values with 95% CI | p-value (paired t-test) |
| --- | --- | --- | --- | --- | --- |
| Full cohort | n | 160 | 160 | 12908.5 | <0.001 |
|  | Average | 17798.5 | 4890 | [ 9833.5 ; 15983.5 ] |  |
|  | CO | 23304.74 | 11945 |  |  |
|  | Minimum | 0 | 0 |  |  |
|  | First quartile | 0 | 0 |  |  |
|  | Median | 12635 | 0 |  |  |
|  | Third quartile | 27900 | 0 |  |  |
|  | Maximum | 91095 | 52420 |  |  |
|  | % of zero values | 46.88 | 79.38 |  |  |
| Cohort with all detected findings | n | 81 | 81 | 24298.4 | <0.001 |
|  | Average | 33503.06 | 9204.71 | [ 19701.9 ; 28894.8 ] |  |
|  | CO | 22262.46 | 15162.13 |  |  |
|  | Minimum | 3900 | 0 |  |  |
|  | First quartile | 14395 | 0 |  |  |
|  | Median | 27900 | 0 |  |  |
|  | Third quartile | 51995 | 14395 |  |  |
|  | Maximum | 91095 | 52420 |  |  |
|  | % of zero values |  | 61.18 |  |  |
| Cohort with significant findings | n | 32 | 32 | 17104.7 | <0.001 |
|  | Average | 24089.22 | 6984.53 | [ 8726.8 ; 25482.6 ] |  |
|  | CO | 15568.37 | 13799.84 |  |  |
|  | Minimum | 7600 | 0 |  |  |
|  | First quartile | 14395 | 0 |  |  |
|  | Median | 19700 | 0 |  |  |
|  | Third quartile | 27900 | 4825 |  |  |
|  | Maximum | 69500 | 52420 |  |  |
|  | % of zero values |  | 71.88 |  |  |

The final economic efficiency of using the AI algorithm in the clinic is shown in Figure 5.

**Figure 5.**
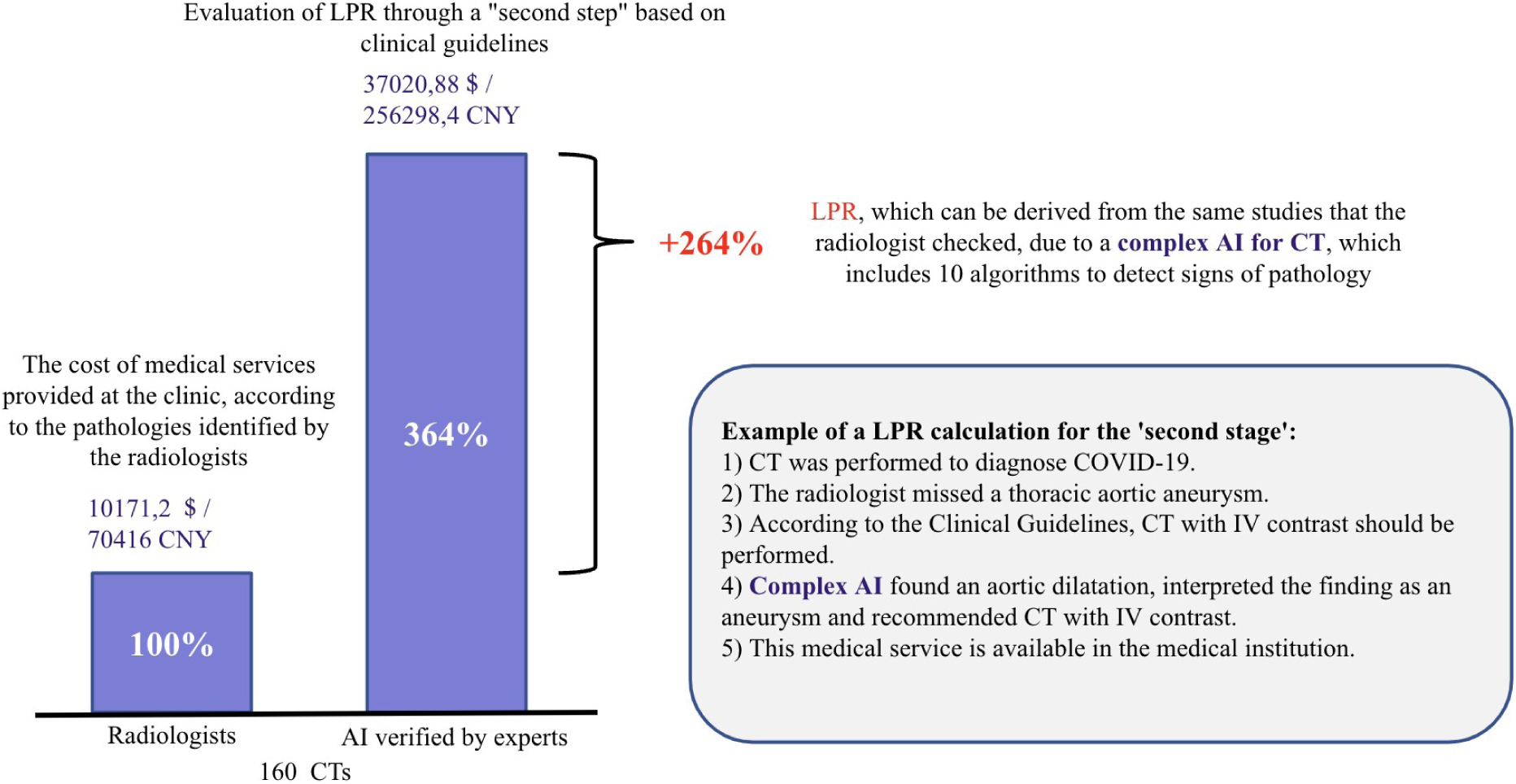
LPR from the use of chest CT complex AI in a healthcare organization. *AI - artificial intelligence; CT - computed tomography; LPR - lost potential revenue*

## An example of the lost potential revenue (LPR) calculation

A clinic’s radiologist correctly described in the report the pulmonary trunk dilation up to 34 mm, an increased Agatston score of up to 350, and a decrease in vertebral density to a maximum of +90 HU. The AI algorithm also detected these pathologies. In addition, the AI found other pathologies - a lung node with a diameter of up to 10*9mm, dilatation of the thoracic aorta up to 45 mm, and thickening of the adrenal gland up to 14 mm. An example of the LPR calculation for this case is presented in Table 8.

**Table 8.** Example of LPR calculation based on data from a single chest CT. *LPR - lost potential revenue*

| Recommendations and costs for all existing pathologies |  |  |  |  |  |  |
| --- | --- | --- | --- | --- | --- | --- |
| Consultations (all) | Further examinations (all) | Follow up (all) | Total cost (all) | Consultations costs (all) | Further examinations costs (all) | Follow up costs (all) |
| Consultations with a cardiologist, endocrinologist | Stress EKG, alkaline phosphatase activity, blood count, calcium levels, BP ultrasound, coronary CT scan, stress ECHO, adrenal tumor hormone study, EchoCG, blood chemistry, CT scan with IV contrast | CT scan, consultation with endocrinologist | 1 191,6 \$/<br>8,195.98 CNY | 102,03 \$/<br>701.78 CNY | 939,14 \$/<br>6,459.52 CNY | 150,43 \$/<br>1,034.68 CNY |
| Recommendations and costs only for pathologies that have been missed by radiologists and found by the algorithm |  |  |  |  |  |  |
| Consultations | Further examinations | Follow up | Total cost | Consultations costs | Further examinations costs | Follow up costs |
| | Stress ECG, coronary CT, stress Echo, adrenal tumor hormone study, EchoCG, CT with IV contrast | CT scan over time | 854,18 \$/<br>5 875,16 CNY | 0 | 754,76 \$/<br>5,191.37 CNY | 99,41 \$/<br>683.79 CNY |

| Recommendations and costs only for pathologies found by radiologists |  |  |  |  |  |  |
| --- | --- | --- | --- | --- | --- | --- |
| Consultations | Further examinations | Follow up | Total cost | Consultations costs | Further examination costs | Follow up costs |
| Consultations with a cardiologist, endocrinologist | Alkaline phosphatase activity test, general blood count, calcium test, abdominal ultrasound, blood chemistry | Consultation with an endocrinologist | 337,42 \$/<br>2,320.82 CNY | 102,03 \$/<br>701.78 CNY | 184,37 \$/<br>1,268.15 CNY | 51,02 \$/<br>350.89 CNY |

## Additional Research Findings

The final results on the number of protocols with significant and non-significant missed pathologies and the percentage of erroneous reports are presented in Table 9. There could be significant and insignificant missed findings in the same report. In total, out of 160 analyzed reports, significant and insignificant findings were detected in 81, which accounted for 50.6% of the total number of CT scans. The average percentage of reports with significant missed pathologies was 28.1% (max 56.9%; min 5%), while for insignificant findings - 27.2% (max 74.1%; min 5%).

**Table 9.**
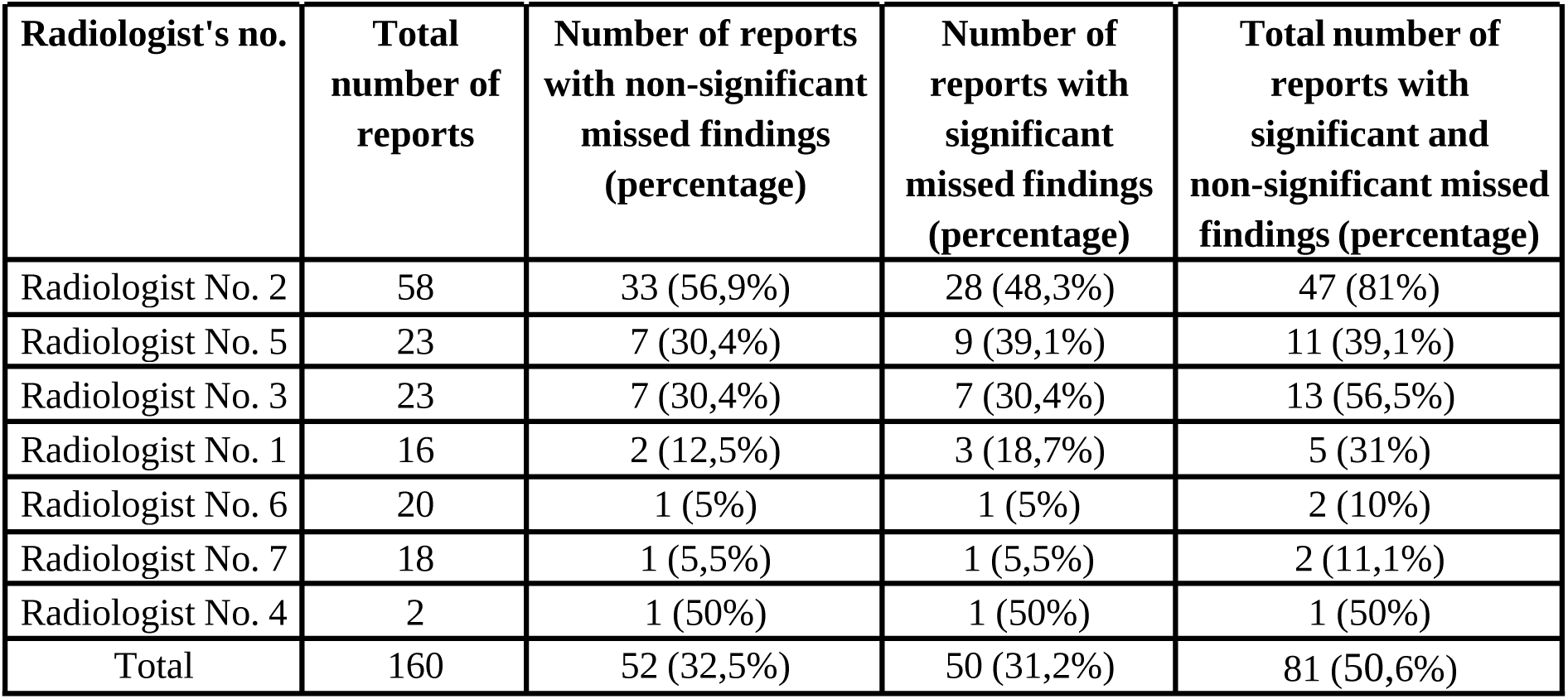
Final results for the number of protocols with critical and non-critical misses.

| Radiologist's no. | Total number of reports | Number of reports with non-significant missed findings (percentage) | Number of reports with significant missed findings (percentage) | Total number of reports with significant and non-significant missed findings (percentage) |
| --- | --- | --- | --- | --- |
| Radiologist No. 2 | 58 | 33 (56,9%) | 28 (48,3%) | 47 (81%) |
| Radiologist No. 5 | 23 | 7 (30,4%) | 9 (39,1%) | 11 (39,1%) |
| Radiologist No. 3 | 23 | 7 (30,4%) | 7 (30,4%) | 13 (56,5%) |
| Radiologist No. 1 | 16 | 2 (12,5%) | 3 (18,7%) | 5 (31%) |
| Radiologist No. 6 | 20 | 1 (5%) | 1 (5%) | 2 (10%) |
| Radiologist No. 7 | 18 | 1 (5,5%) | 1 (5,5%) | 2 (11,1%) |
| Radiologist No. 4 | 2 | 1 (50%) | 1 (50%) | 1 (50%) |
| Total | 160 | 52 (32,5%) | 50 (31,2%) | 81 (50,6%) |

The total number of reports statistically significantly increased the number of errors. The total work experience in radiology (excluding residency) and thoracic radiology (including residency) reduced the number of mistakes. However, these data are not representative due to the small sample of radiologists and the presence of a dominant case. Detailed data on the experience of doctors are presented in supplementary materials.

Examples of the AI algorithm work are shown in Figures 6 and 7.

**Figure 6.**
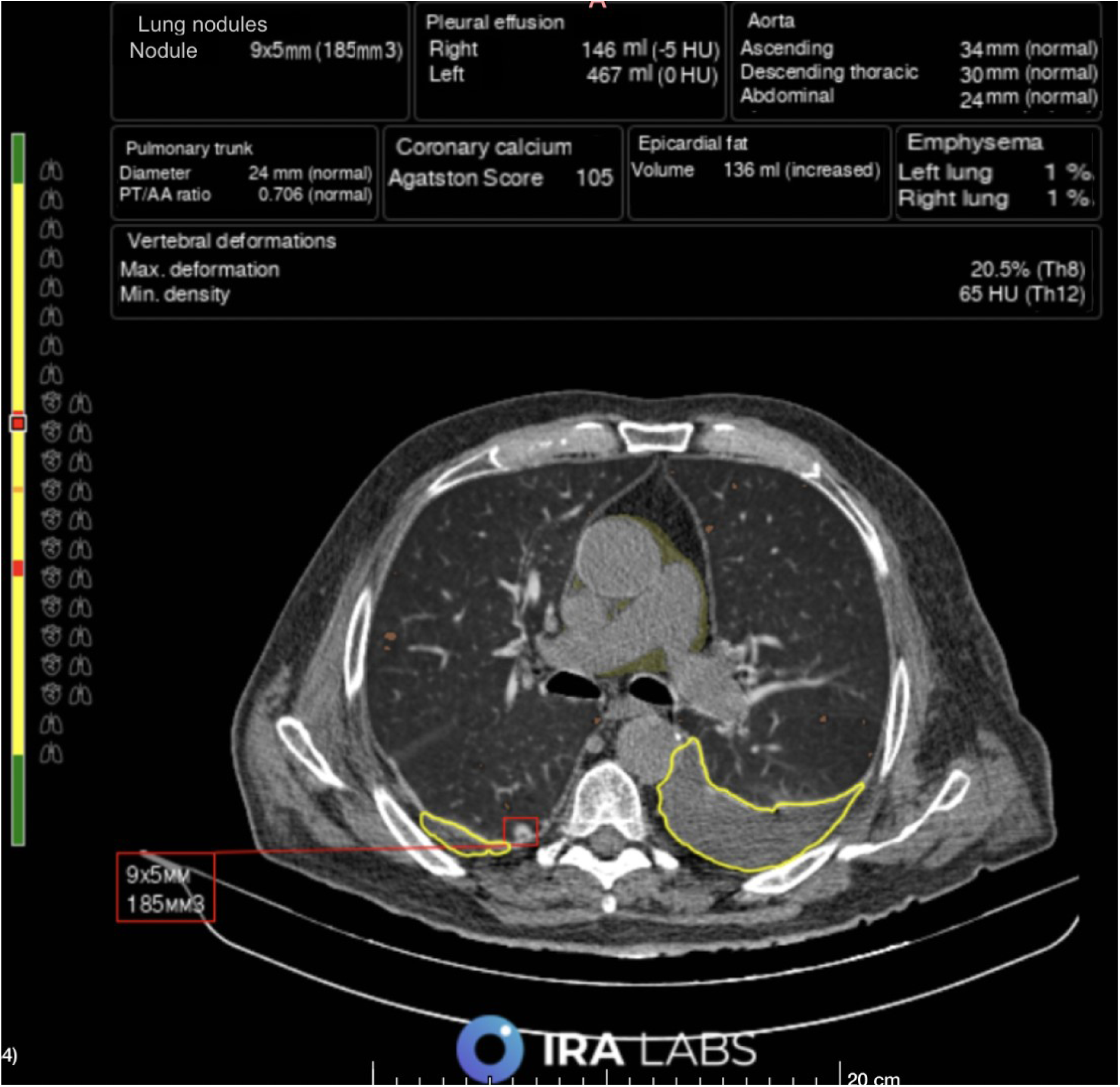
Patient A. The radiologist correctly identified bilateral hydrothorax and emphysematous changes but did not describe a pulmonary nodule in the right lung. The AI algorithm detected all three pathologies: hydrothorax is outlined with a yellow line, emphysematous changes are outlined with orange, and lung nodule is outlined with a red square.

**Figure 7.**
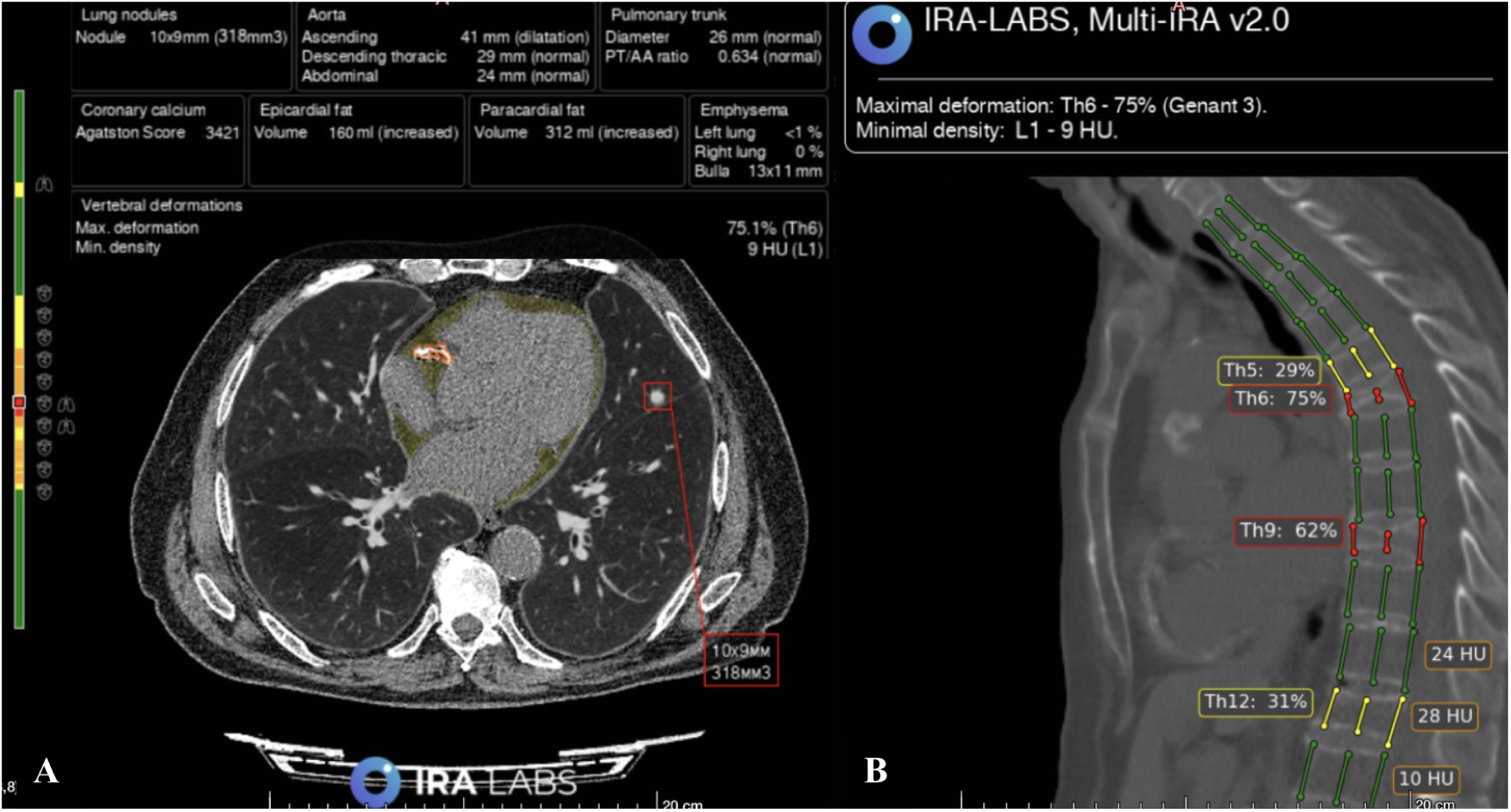
Patient B.. A is an axial CT scan. The radiologist and algorithm correctly identified a lung nodule in the left lung (marked with a red square) and coronary artery calcification (outlined with an orange line). Additionally, the algorithm indicated increased epicardial fat (filled in yellow, this pathology was not considered in the study). B - sagittal CT scan. The radiologist and algorithm correctly identified compression fractures of Th6 and Th9 vertebral bodies, Genant 3 (3 columns are marked with red lines). However, the radiologist did not indicate deformities of Th5 and Th12 vertebral bodies, Genant 2 (3 columns marked with yellow lines) in the report.

## Adverse events

As a result of the study, no adverse events were noted.

## Discussion

### Summary of the primary results of the study

As a result of the study, for the first time, it was possible to show the expected economic effect that can be obtained using a complex AI product for chest CT analysis.

Medical services would need to be provided to patients following current clinical guidelines. The total LPR of the “second stage” of the necessary diagnostics only for those pathologies missed by radiologists, but identified by AI, amounted to just over 2 million rubles, or 3.6 times more than the cost of medical services based on the pathologies reported by radiologists (without AI usage). While the LPR only from significant missed pathologies amounted to slightly more than 770 thousand rubles, according to the clinic price list, or 98% more than the cost of medical services that can be provided in the clinic following the pathologies identified by radiologists (without AI usage).

The possibility of using a complex AI product for auditing CT radiology reports is also shown.

### Discussion of the main result of the study

An analysis of the socioeconomic burden of the COVID-19 pandemic can serve as an example of the significant social and economic consequences of a mass disease for the Russian healthcare system and society as a whole, which focuses attention not only on the clinical but also on the economic importance of investing in the development of strategies to combat diseases [43]. According to experts, the socioeconomic burden of COVID-19 in the Russian Federation in 2020 amounted to about 5.4 trillion rubles (5% of the nominal GDP in 2020), which corresponds to 2,486 YLL (years of life lost; years of life lost due to premature mortality of the population) among men and 1,378 YLL among women [43]. The economic burden of non-communicable diseases in the Russian Federation for the same year amounted to 4 trillion rubles. At the same time, the damage from chronic diseases is comparable to the budget for the entire health care of the Russian Federation, and the funds that can be released through effective prevention could become a substantial additional resource for the country’s development [44].

In the available literature, we did not find studies evaluating the impact of complex AI systems for the analysis of CT scans on the economic aspects of the work of a medical organization.

Pickhardt et al. built a model of the cost and clinical effectiveness of screening based on complex AI analysis of abdominal CT studies [45]. Using expected disease prevalence, transition probabilities between health conditions, associated health care costs, and treatment efficacy for three diseases (cardiovascular disease (CVD), osteoporosis, and sarcopenia), three mutually exclusive screening models were assessed: (1) ignoring results ("not treat"; no intervention regardless of CT findings), (2) universal statin therapy ("treat all" for CVD prevention without consideration of CT findings), and (3) opportunistic screening for CVD, osteoporosis, and sarcopenia with abdominal CT scans based on AI analysis (targeted treatment of individuals at risk). For baseline scenarios for groups of 55-year-old men and women simulated over ten years, AI-assisted CT-based opportunistic screening was a cost-effective and more effective clinical strategy than "ignore" and "treat all" approaches. The authors summarize that AI-assisted CT-based opportunistic screening appears to be a highly cost-effective and clinically effective strategy under a wide range of input assumptions and provides cost savings in most scenarios. However, compared to our study, a real working complex AI was not presented. In addition, our work used complex AI targeting ten target pathologies compared to three in the referenced paper. In this regard, there is hope for an increase in the potential for the cost-effectiveness of the AI systems application in combination with its positive impact on diagnostics quality.

Few publications have examined the economic impact of such programs using an integrated approach for several pathologies without using AI systems. Thus, for the Netherlands, it was calculated that comprehensive LDCT screening for three diseases, such as lung cancer, chronic obstructive pulmonary disease (COPD), and CVD in people aged 50 to 75 years, could be cost-effective if it costs less than 971 euros per person examined [46]. The estimated maximum acceptable cost per person screened for lung cancer with LDCT was €113 (for the Netherlands). In addition, experts estimated the additional charges required for examining patients with incidental extrapulmonary findings [47]. The total cost of treating all incidental findings based on the results of this study was $ 26,321, and the average price of diagnostic testing for each malignancy was $ 1,316. The most expensive was the verification of cases of thyroid cancer. According to the authors, analyzing the costs of additional diagnostic and therapeutic measures associated with extrapulmonary changes detected during lung cancer LDCT screening is one of the main stages in proving the cost-effectiveness of such actions. This approach (although limited by SRL using chest CT) allows us to propose using complex AI to improve the diagnostic and cost-effectiveness of studies.

Our study also used complex AI to search for ten pathologies. Such analyses, referred to as stock analysis, have proven helpful in making informed decisions about further research [48-50]. They are preferred during the diagnostic intervention and evidence development to optimize data collection and more accurately assess long-term health economic impact when a large amount of clinical data becomes available.

Before the COVID-19 pandemic, AI algorithms were used to detect radiographic symptoms for disease detection, classification, image optimization, radiation dose reduction, and workflow improvement [51]. Medical research on the effectiveness of AI makes such programs more understandable, safer, more efficient, and integrated into doctors’ workflows [52]. Currently, studies are underway within the IMALife project, studying the reduction of mortality not only from lung cancer but also from the consequences of emphysema (COPD biomarker) and coronary calcification (atherosclerosis biomarker) [53].

Evaluation of the effectiveness of the use of AI has so far only been found in AI algorithms designed to search for only one target pathology. In a study by Ziegelmayer and colleagues, the base scenario CT + AI showed a negative incremental cost-effectiveness ratio (ICER) compared to CT, demonstrating lower costs and higher efficiency. Threshold analysis showed that ICER remained negative until the $68 threshold to support AI applications. Thus, using a mono system for analyzing LDCT data using AI for SRL is a reasonable diagnostic strategy for cost-effectiveness [54].

The constantly growing volume of radiological examinations creates an additional burden on the radiologist [55]. The excessive workload can increase the likelihood of errors and jeopardize the quality of care [56]. The audit system with a retrospective double review of studies is widely used in radiology. The best known is the "RADPEER system" of the American Society of Radiologists [57]. However, according to a study by Lauritzen et al., double-reading one-third of the studies performed at their institution takes up about 20-25% of physicians’ working time [58]. Using AI algorithms can significantly reduce the time to review studies and increase the scope and quality of audits. The use of AI also affected the quality of work of radiologists in the form of a change in the ratio of the severity of lung lesions with suspected COVID-19 towards a reduction in the proportion of the severe and critical assessment of the severity of lung lesions [59].

In our study, we also demonstrated the feasibility of using complex AI to audit radiology reports. We identified more than 28% of reports with significant and 27% of protocols with non-significant missed pathologies. It should be noted that for all CT examinations, radiologists described the main diagnostic tasks (pathologies) for which patients were sent for scanning. The clinic’s radiologists did not have tools at all workstations for quick measurement of the Agatston index. At the same time, the measurement of the density of the vertebral bodies was not included in the standard for studies reporting in this radiology department. In addition, it has been shown that the average error rate is comparable between doctors, so firing 1-2 radiologists with the worst results according to audits, the clinic will not solve the problem of missing pathologies.

In this paper, when calculating the economic effect, the AI costs, as well as the cost of work performed by experts in validating AI results, were not taken into account. The cost of these expenses is variable depending on the number of algorithms, the level of experts involved, and other factors. Any tariff for an integrated AI service will be cost-effective, provided that the total costs are lower than the profit of a medical organization, thanks to AI usage (Figure 8). However, such an analysis is beyond the scope of the current study.

**Figure 8.**
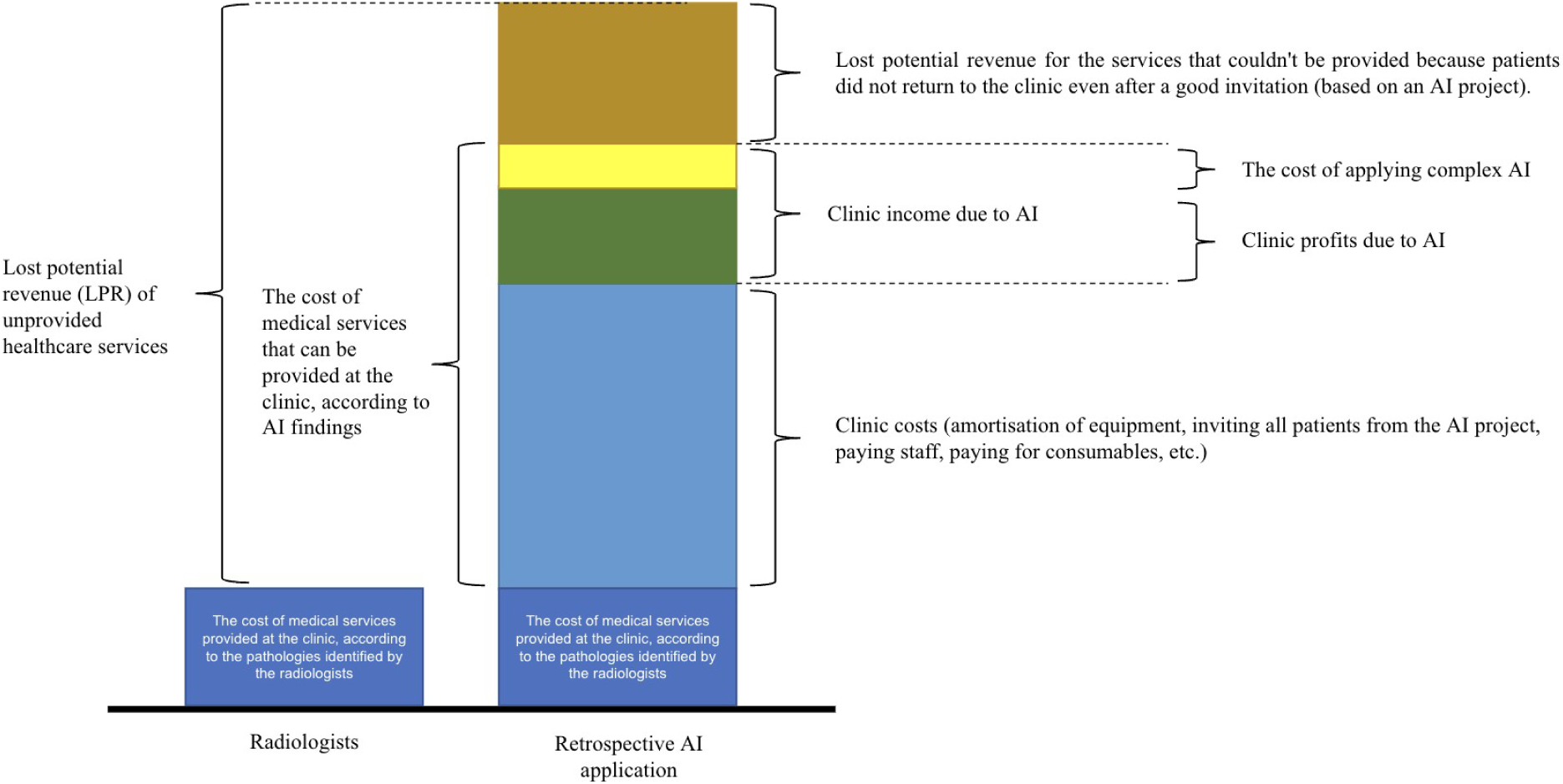
Lost potential revenue (LPR) due to the lack of application of complex AI for CT in a medical organization, taking into account the cost of AI usage. *AI - artificial intelligence*.

### Study Limitations

This study is a pilot study and has several limitations. It has a retrospective design and evaluates the maximum potential of a private healthcare organization based on recommendations following pathological findings. In practice, not all patients are inclined to listen to doctors and follow reasonable recommendations, especially when it comes to paid additional examinations and consultations. In addition, conversion in each clinic has its own characteristics that go beyond the current study’s design.

The purpose of this study was only to estimate the cost of the "second stage" without the cost of the "third" and subsequent stages; that is, the cost of treatment and rehabilitation was not considered. However, for almost every pathology that complex AI can detect, the cost of treatment significantly exceeds the cost of the "second stage". In every medical organization, many factors affect the quality of the work of doctors. The indicator of the quality of radiologists’ work (the number of missed clinically significant radiographic findings) can be variable depending on the experience of radiologists, the number of studies per day, time of day, day of the week, and many other additional indicators that can affect the knowledge, attentiveness, and readiness of the radiologist describe all pathological findings and reasonable recommendations for the "second stage" in the protocol.

In our work, the potential false negative AI findings were not assessed since the AI system was previously validated during independent testing on the non-public datasets of the Moscow experiment. The selected AI settings were found to be acceptable and calibrated for work.

The purpose of this study was not to assess the economic effect at the city and federal healthcare levels. However, each medical organization in the Russian Federation is not deprived of the opportunity to provide paid medical services to the population, justified in terms of the principles of evidence-based medicine.

Our study did not examine patients’ compliance with invitations based on a retrospective analysis. At the same time, the revealed findings are closely related to the time intervals between the CT examination and the "second stage".

## Conclusion

Using a complex AI system to analyze chest CT data as an assistant to the radiologist to diagnose ten common and important pathological findings leads to improved detection. It is essential that LPR with this approach is 3.6 times larger compared to the standard model of work of radiologists without AU usage. Opportunistic screening of multiple diseases requires a detailed study of comorbidities to determine the optimal target group for diagnostic intervention using complex AI. The use of complex AI for chest CT will likely be cost-effective since this approach reveals many significant pathological changes that require additional medical services.

## Additional Information

### Financial source

The study was carried out at the expense of the authors’ funds.

### Conflict of interest

The authors declare no apparent potential conflicts of interest related to the publication of this article.

### Author participation

The authors confirm that their authorship complies with the international ICMJE criteria (all authors contributed significantly to developing the concept, research, and preparation of the article and read and approved the final version before publication).

### The most significant contribution is distributed as follows

Chernina V.Yu. — search for publications on the article’s topic, formation of a data set, writing the text of the manuscript;

Belyaev M.G. — expert evaluation of the data, manuscript’s text editing;

Silin A.Yu. — the concept of the study;

Avetisov I.O. — manuscript’s text editing;

Pyatnitskiy I.A. — manuscript’s text editing; English text translation;

Petrash E.A. — manuscript’s text editing;

Basova M.V. — formation of a data set, processing of research results;

Sinitsyn V.E. — expert evaluation of the data, manuscript’s text editing;

Omelyanovsky V.V. — expert evaluation data;

Gombolevsky V.A. — the concept of the study, expert evaluation of data, manuscript’s text writing, approval of the final version.

## Data Availability

All data produced in the present study are available upon reasonable request to the authors.

## Acknowledgments

The team of authors is grateful to Pisov M.E., Technical Director of IRA Labs, and the development team (Proskurov V.A., Samoylenko A.I., Borzov A.S., Bukharaev A.N., Goncharov M.Yu., Shirokikh B.N., Kurmukov A.I., Nachinkin I.A., Telepov A.Yu., Shimovolos S.A., Donskovoy M.A., Samokhin V.Yu., Shevtsova A.E., Yaushev F., Tkachenko S.M., Zakharov A.A., Saparov T.N., Filipenko V.K., Tominin V.D., Tominin Ya.D., Samoilenko A.I., Malevanny V.M., Leonov A.Yu., Nogina D.S., Bazarova A.I., Marakhovsky K.V., Belkov A.S., Vasin A.A., Berezhnoy D.S., Musikhin M.M.); Dugova M.N., medical director, and a team of medical experts (Ilyicheva D.V., Sevryukov D.D., Shchipakhina Ya.A., Lyubimaya Yu.O., Aleshina O.O., Tsybulskaya Yu.A.); Lamzin M.S., Operations Director, Gareeva R.R., Product Manager of the company, Panina E.V., Deputy Director for Business Development, and all the staff at the Yauza Hospital.

## List of abbreviations

AI: - artificial intelligence
CI: - confidence interval
COPD: - chronic obstructive pulmonary disease
CT: - computed tomography
CVD: - cardiovascular disease
Dataset: - a set of data, a collection of logical records
ICD-10: - International Classification of Diseases 10th revision
LCS: - Lung Cancer Screening
LDCT: - low dose computed tomography
LPR: - lost potential revenue
MILD: - Multicentric Italian Lung Detection
MRI: - magnetic resonance imaging
NLST: - National Lung Screening Trial
PACS-RIS: - Picture Archiving and Communication System Radiology information systems

## Supplementary materials

The experience of radiologists is indicated in the table.

**Table.**
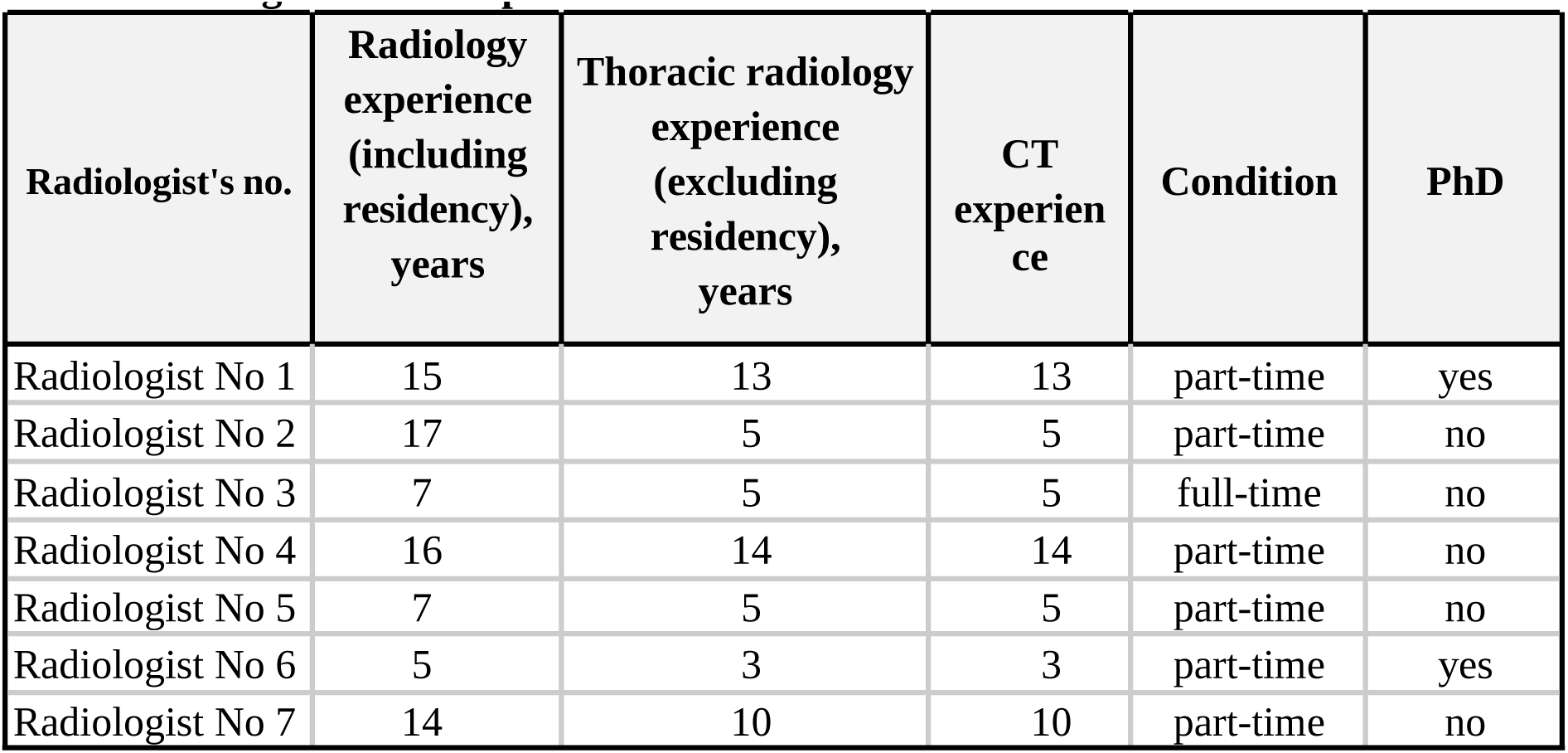
Radiologists’ work experience.

### Generalized Linear Model

The generalized linear model assumes that each observation Yi depends linearly on the values of the variables Xip, p = 1,2,…m:

Yi = bi0 + b1Xi1 + b2Xi2 + … + bmXim + εi

, where the variables X can be either categorical, taking into account groups, classes, categories, or continuous. In the generalized linear model, the coefficients {bj, j = 0,1,2,…,m} are estimated for the model of the dependence of the parameter Y on the factors {Xj, j = 1,2,…,m}. These coefficients’ statistical significance is determined (calculation of p-values for testing hypotheses Hj0: bj = 0, j = 0,1,2,…,m), which shows the significance of the influence of the corresponding factors on the target parameter.

In our case, some parameters have a lognormal distribution, so a logarithmic transformation has been applied to them. As a result, the model looks like this:

Y = log(Total number of radiology reports with significant and non-significant errors)

X1 = All radiology reports

X2 = log (Work experience in thoracic radiology (excluding residency))

X3 = log (Length of experience in CT)

And the categorical parameter X4 = Academic degree

The evaluation results of the OLM coefficients and their statistical significance are shown in the table.

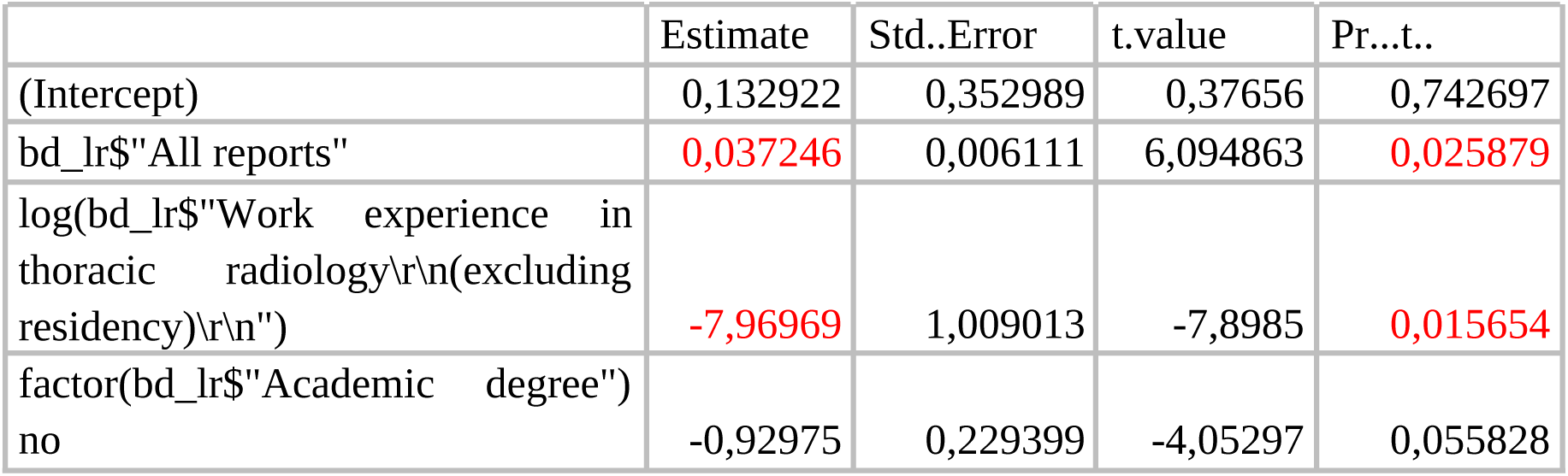

It can be concluded that the total number of radiology reports statistically significantly increases the number of errors. The combined work experience in radiology and thoracic radiology reduces the number of errors. However, these data are not representative due to the small sample of radiologists and the presence of a dominant case.

Partially similar results (up to statistical significance and the influence of one of the types of experience) are also reflected in the correlation analysis:

ρ(log(Total number of protocols with critical and non-critical errors), All protocols) = 0.88 with 95% CI [0.39; 0.98] and p-value = 0.008 (correlation is statistically significant) ρ(log(Total number of reports with significant and non-significant errors), log(Work experience in thoracic radiology (excluding residency))) = -0.45 with 95% CI [-0.9; 0.46] and p-value = 0.31 (correlation is not statistically significant)

ρ(log(Total number of reports with significant and non-significant errors), log(Work experience in thoracic radiology (excluding residency))) = -0.27 with 95% CI [-0.85 0.61], and p-value = 0.56 (correlation is not statistically significant)

However, without the statistical significance of correlations, it is impossible to talk about any trends.

## Notes

### Competing Interest Statement

The authors have declared no competing interest.

### Funding Statement

This study did not receive any funding.

### Author Declarations

The Independent Ethics Committee of the Moscow Regional Branch of the Russian Society of Roentgenologists and Radiologists gave ethical approval for this work on 01.03.2023.

### Summary of Updates

Figure 1 revised; added figure 2.

