## Supplementary materials for "A diagnostic and economic evaluation of the complex artificial intelligence algorithm aimed to detect 10 pathologies on the chest CT images"

The experience of radiologists is indicated in the table.

**Table. Radiologists' work experience.**

| **Radiologist's no.** | **Radiology experience (including residency),**  **years** | **Thoracic radiology experience**  **(excluding residency),**  **years** | **CT experience** | **Condition** | **PhD** |
| --- | --- | --- | --- | --- | --- |
| Radiologist No 1 | 15 | 13 | 13 | part-time | yes |
| Radiologist No 2 | 17 | 5 | 5 | part-time | no |
| Radiologist No 3 | 7 | 5 | 5 | full-time | no |
| Radiologist No 4 | 16 | 14 | 14 | part-time | no |
| Radiologist No 5 | 7 | 5 | 5 | part-time | no |
| Radiologist No 6 | 5 | 3 | 3 | part-time | yes |
| Radiologist No 7 | 14 | 10 | 10 | part-time | no |

**Generalized Linear Model**

The generalized linear model assumes that each observation Yi depends linearly on the values of the variables Xip, p = 1,2,...m:

Yi = bi0 + b1Xi1 + b2Xi2 + ... + bmXim + εi

, where the variables X can be either categorical, taking into account groups, classes, categories, or continuous. In the generalized linear model, the coefficients {bj, j = 0,1,2,...,m} are estimated for the model of the dependence of the parameter Y on the factors {Xj, j = 1,2,...,m}. These coefficients' statistical significance is determined (calculation of p-values for testing hypotheses Hj0: bj = 0, j = 0,1,2,...,m), which shows the significance of the influence of the corresponding factors on the target parameter.

In our case, some parameters have a lognormal distribution, so a logarithmic transformation has been applied to them. As a result, the model looks like this:

Y = log(Total number of radiology reports with significant and non-significant errors)

X1 = All radiology reports

X2 = log (Work experience in thoracic radiology (excluding residency))

X3 = log (Length of experience in CT)

And the categorical parameter X4 = Academic degree

The evaluation results of the OLM coefficients and their statistical significance are shown in the table.

|  | Estimate | Std..Error | t.value | Pr...t.. |
| --- | --- | --- | --- | --- |
| (Intercept) | 0,132922 | 0,352989 | 0,37656 | 0,742697 |
| bd_lr$"All reports" | 0,037246 | 0,006111 | 6,094863 | 0,025879 |
| log(bd_lr$"Work experience in thoracic radiology\r\n(excluding residency)\r\n") | -7,96969 | 1,009013 | -7,8985 | 0,015654 |
| factor(bd_lr$"Academic degree") no | -0,92975 | 0,229399 | -4,05297 | 0,055828 |

It can be concluded that the total number of radiology reports statistically significantly increases the number of errors. The combined work experience in radiology and thoracic radiology reduces the number of errors. However, these data are not representative due to the small sample of radiologists and the presence of a dominant case.

Partially similar results (up to statistical significance and the influence of one of the types of experience) are also reflected in the correlation analysis:

ρ(log(Total number of protocols with critical and non-critical errors), All protocols) = 0.88 with 95% CI [0.39; 0.98] and p-value = 0.008 (correlation is statistically significant)

ρ(log(Total number of reports with significant and non-significant errors), log(Work experience in thoracic radiology (excluding residency))) = -0.45 with 95% CI [-0.9; 0.46] and p-value = 0.31 (correlation is not statistically significant)

ρ(log(Total number of reports with significant and non-significant errors), log(Work experience in thoracic radiology (excluding residency))) = -0.27 with 95% CI [-0.85 0.61], and p-value = 0.56 (correlation is not statistically significant)

However, without the statistical significance of correlations, it is impossible to talk about any trends.
